# Prevalence of Olfactory Dysfunction with the Omicron Variant of SARS-CoV-2: A Systematic Review and Meta-analysis

**DOI:** 10.1101/2022.12.16.22283582

**Authors:** Christopher S. von Bartheld, Lingchen Wang

## Abstract

The omicron variant is thought to cause less olfactory dysfunction than previous variants of SARS-CoV-2, but the reported prevalence differs greatly between populations and studies. Our systematic review and meta-analysis provide information about regional differences in prevalence as well as an estimate of the global prevalence of olfactory dysfunction based on 62 studies reporting on 626,035 patients infected with the omicron variant. Our estimate of the omicron-induced prevalence of olfactory dysfunction in populations of European ancestry is 11.7%, while it is significantly lower in all other populations, ranging between 1.9% and 4.9%. When ethnic differences and population sizes are taken into account, the global prevalence of omicron-induced olfactory dysfunction in adults is estimated at 3.7%. Omicron’s effect on olfaction is twofold to tenfold lower than that of the alpha or delta variant, according to previous meta-analyses and our analysis of studies that directly compared prevalence of olfactory dysfunction between omicron and previous variants. The profile of prevalence differences between ethnicities mirrors the results of a recent genome-wide association study that implicated a gene locus encoding an odorant-metabolizing enzyme, UDP glycosyltransferase, to be linked to the extent of COVID-related loss of smell. Our analysis is consistent with the hypothesis that this enzyme contributes to the observed population differences.

## 1. Introduction

The omicron variant has been reported to cause less anosmia than the preceding SARS-CoV-2 virus variants [1-4]. Since the prevalence of olfactory dysfunction varies greatly between studies, the global prevalence of anosmia caused by omicron has not yet been estimated. The number of confirmed COVID-19 cases reported to the World Health Organization (WHO) by November 30, 2022 was 639 million (WHO Coronavirus (COVID-19) Dashboard, https://covid19.who.int/), but the true number of cases is believed to be much higher, at about 3.4 billion in October 2021 [5]. A total of 6 billion cases – after the global spread of the more infectious omicron variant – has been estimated in October 2022 [6], Since the prevalence of olfactory dysfunction differs between virus variants [1-4,7,8], it is important for estimates of the current global and regional prevalence of olfactory dysfunction to take properties of different virus variants into account. It has been argued that, even though omicron may cause a lower prevalence of olfactory dysfunction, the increased infectivity may produce equivalency or even a net gain in the cases of hyposmia or anosmia, because a much larger number of people will become infected with the omicron variant [9,10].

It is possible that host factors also contribute to the population differences in COVID’s olfactory dysfunction [7,11-13]. Such host factors, besides age and gender, are apparently not due to differences in expression levels or in genetic variation of the virus entry proteins, ACE2 and TMPRSS2, as was initially assumed [11,12,14], but rather may be due to genetic variation and the frequency of risk alleles of an odorant-metabolizing enzyme, a glycosyltransferase that is encoded by the UGT2A1/A2 locus [13]. This enzyme is abundantly expressed in sustentacular support cells of the olfactory epithelium of vertebrates [15,16], including humans [17-20].

Here, we conducted a systematic review and meta-analysis of the literature on olfactory dysfunction caused by the omicron variant. In this review, we focused on loss of smell rather than loss of taste. Loss of taste is thought to be, in part, due to loss of smell [21], and therefore we grouped the diverse reports on “loss of smell”, “loss of smell and taste”, and “loss of smell or taste” in one single category. Furthermore, because the large majority of reports use patients’ subjective recall to identify new olfactory dysfunction, we restricted our analysis to studies that used subjective methodology (the patient’s recollection of changes in smell), rather than objective psychophysical testing, which depends on cultural context and therefore requires population-specific validation [22]. For omicron-associated olfactory dysfunction, we found 62 studies reporting on the basis of the patient’s subjective recall, and only one study that also performed psychophysical testing.

We generated estimates of the global prevalence of omicron-induced olfactory dysfunction, as well as regional prevalence, which is determined at least in part by genetics (prevailing ethnicity) within populations. Similarities between the results of our analysis and those of a recent genome-wide association study [13] point to differences in the frequency of the risk allele for an odorant-metabolizing enzyme as a contributing factor, resulting in population differences in the prevalence of olfactory dysfunction.

## 2. Methods

### 2.1 Search Strategy

For our systematic review of the literature, we adhered to the guidelines set forth by the Preferred Reporting Items for Systematic Reviews and Meta-Analyses (PRISMA) [23]. Reports of studies that estimate the prevalence of olfactory dysfunction were identified through a search of three databases, PubMed, Google Scholar, as well as the iCite NIH COVID portal (https://icite.od.nih.gov/covid19/search/), for the years 2021, 2022 and 2023. The COVID portal was included in order to capture preprints in addition to peer-reviewed articles. The following search strategy was formulated using the keywords “omicron” and “smell”, as well as “omicron” and “anosmia.” Only English terms were used for the search strategy. Reference lists from the eligible articles were examined to identify additional relevant studies. Duplicates were removed, but the first date of publication, in case of preprints, was recorded, even when the peer-reviewed version of the paper, when available, was compiled in our list of references. All titles were screened, and when potentially relevant, the abstract was evaluated to decide whether the paper should be short-listed for full-text reading. The full text of all short-listed records was reviewed to determine whether they were eligible according to our inclusion and exclusion criteria, and then were used to produce the final selection of studies for inclusion in subsequent analyses (Fig. 1).

**Figure 1.**
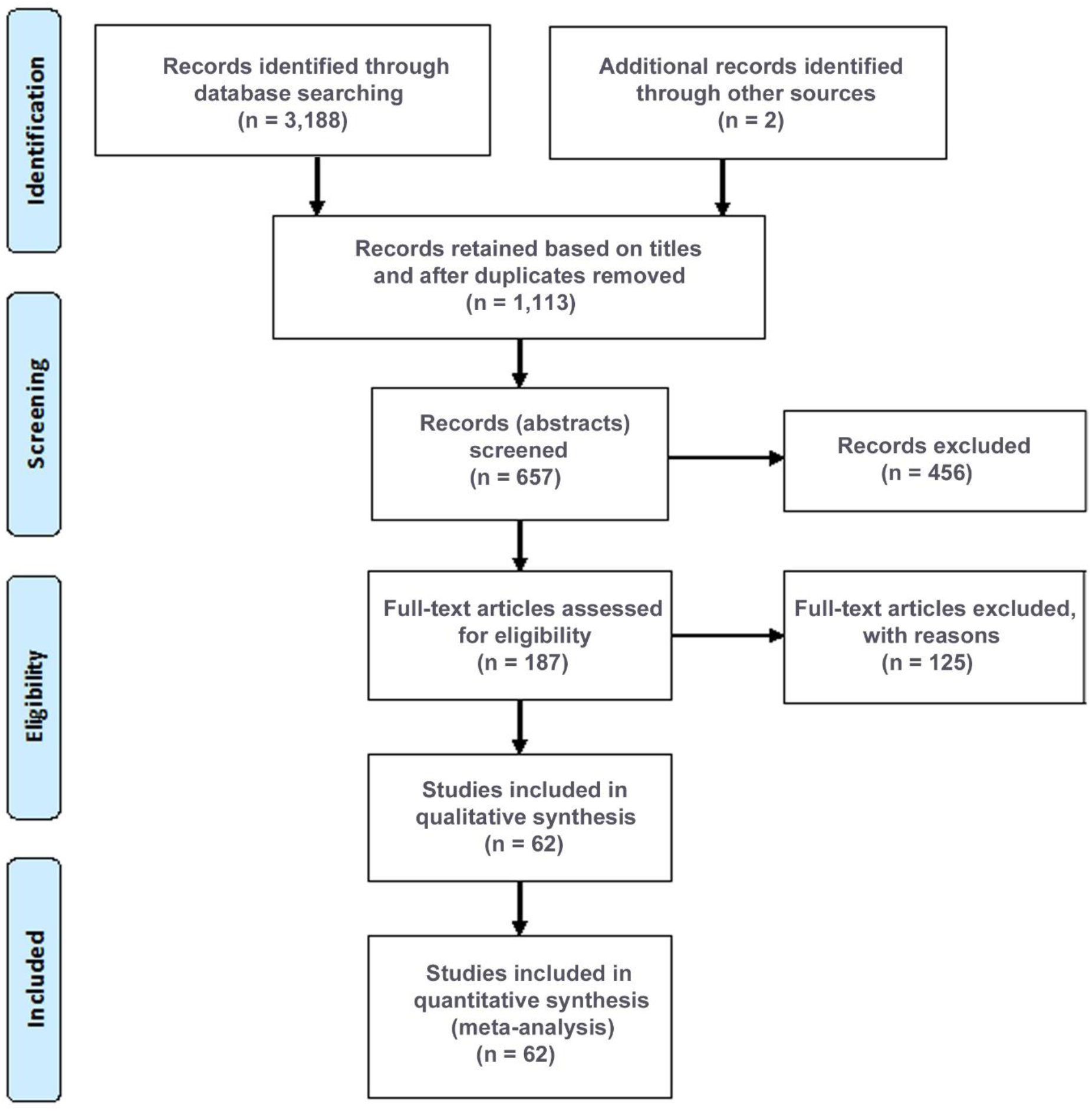
Flowchart illustrating the Literature Search, Systematic Review and Meta-analysis according to the PRISMA guidelines. PubMed, the NIH COVID portal, and Google Scholar were searched sytematically; other sources were found in references of articles and in news media. The literature was last updated on January 10, 2023. The reasons for exclusion of full-text articles were, in decreasing frequency: no prevalence data; no data for omicron; long (not acute) COVID; children only; review; case report.

### 2.2 Inclusion/Exclusion Criteria

Studies which were deemed eligible for the systematic review met all of the following inclusion criteria: (1) studies reporting the numerical prevalence of olfactory dysfunction in humans infected with the omicron variant (B.1.1.529) and any of the omicron subvariants, BA.1, BA.2, BA.1.1, BA.2.2, BA.2.10, BA.2.38, BA.2.75, BA.5, BQ.1, XBB. Report of an odds ratio only was not sufficient for inclusion [24]; (2) studies on adults or adolescents (when a small number of children was included, this was considered acceptable), but studies that focused entirely on children were not included, because it is known that children with COVID have a significantly lower prevalence of olfactory dysfunction than adults with COVID [25]; (3) evidence of infection with SARS-CoV-2; genomic proof of variant type was not deemed necessary when it was known that the vast majority of infections during the period and in the region of data collection were omicron cases rather than cases caused by another virus variant; (4) olfactory dysfunction was monitored through subjective recall, and all members of the cohort were specifically asked about changes in smell, changes in smell or taste, or changes in smell and taste; review of medical records for entries about loss of smell, but without universal and specific questioning of patients, was not acceptable (e.g., [26]); (5) the olfactory dysfunction occurred during the acute phase of infection – long-term studies inquiring about changes of smell persisting for weeks or months after the infection were not included. Comparison with variants other than omicron was not a required inclusion criterion.

### 2.3 Quality Assessment and Publication Bias

Risk of bias in cohort studies was assessed using a modified Newcastle-Ottawa scale (adapted Cochrane’s risk of bias tool [27]). This scale attempts to assess accuracy of measurements, as well as whether the cohort is representative of the community. Duration of follow-up is not relevant for the current review and analysis. It does assess study design and cohort size, as well as information about convenience samples and response rates, when applicable. In addition, we explored the magnitude of the potential bias caused by survey-type studies that rely on the initiative and motivation of respondents [9,28-30] by comparing the results of traditional-design studies with those of survey-type studies. In addition, we generated funnel plots to assess potential publication bias [31].

### 2.4 Data Extraction

The relevant data of each study were extracted by using pre-designed tables, including the first date of publication, first author name, country, geographic region, the cohort size, the number of cases, and the percentage calculated from the number of cases per cohort. When applicable, the comparator virus variant was also noted, along with its cohort size, number of cases with olfactory dysfunction, and the percentage, as well as the name(s) of the previous variant or variants causing the infection. When the comparator virus variant was not disclosed, it was retrieved as G614 vs D614 [32]. Additional information about cohorts such as age, gender, and ethnic composition was recorded when studies provided this information.

### 2.5 Subgroup Analyses and Comparisons

The global prevalence of olfactory dysfunction due to omicron infection was calculated by taking ethnic differences and population sizes into account, and this prevalence was compared with the global and regional prevalence due to previous variants, using information from studies that reported such data (32 out of 62 studies). Because of ethnic differences between populations, the prevalence for each major ethnicity (European ancestry, African, Middle East, East Asian, South Asian, Latino/Hispanic) was estimated separately and weighted by population size to calculate an estimate of the current global hyposmia prevalence due to omicron. This was necessary to prevent bias due to the fact that the largest fraction of available studies and those with the largest cohorts have focused on people with European ancestry.

### 2.6 Data Synthesis

The primary purpose of the meta-analysis was to produce a more precise and reliable estimate of the effect of the omicron variant on olfactory dysfunction, and to compare this estimate with previous estimates that were made pre-omicron (results from previous meta-analyses) as well as preparing a direct comparison by compiling the data from those studies which provided internal comparative data on other virus variants.

### 2.7 Statistical Analyses

Pooled analyses were performed for olfactory dysfunction prevalence and risk ratio (RR). The heterogeneity among studies was evaluated by Cochran’s Q test and the I^2^ index [33,34]. The random-effect models were used to conservatively diminish the heterogeneity between the studies [34]. The study weights were obtained based on the DerSimonian-Laird method [36]. A continuity correction of 0.5 was applied to studies with zero cases [35]. Subgroup pooled analyses were conducted by ethnicity and study type (survey-type studies and traditional-design studies). Meta-regression analyses were performed to test the association between prevalence and key variables [35], including the UGT2A1 risk allele frequency [13] and the study type [36]. The risk of publication bias was evaluated using funnel plots and Egger’s test [37]. The significance level was set to 0.05. All the meta-analyses were performed using the Stata SE 16.0 software (StataCorp, TX, USA).

## 3. Results

### 3.1 Properties of Studies

We found 62 studies, published between November 27, 2021 and January 10, 2023, that met our inclusion criteria. Collectively, these studies reported the olfactory status of 626,035 patients infected with the omicron virus (Table 1). These studies were conducted in 26 countries on six continents (Fig. 2). Twenty-five studies were from populations primarily of European ancestry [9,30,38-60], seventeen studies on East Asians [61-77] (note that one of these studies, [75], reports on the same cohort as [74], and therefore was removed from the meta-analysis), eight studies on South Asians [79-86], four studies on Latinos/Hispanics [87-90], five studies on populations in Africa [91-95], and three studies from the Middle East [96-98]. The location of studies, with the prevalence indicated by the color intensity, and the cohort size indicated by the size of the circles, shows that Western countries report the highest prevalence, while studies from East Asia and the Middle East report the lowest prevalence (Fig. 2). We found that five studies were of low quality, 36 of moderate quality, and 21 of high quality according to the modified Newcastle-Ottawa scale [27]. Thirty-two of the 62 studies also reported the olfactory status caused by one or more of the previous SARS-CoV-2 virus variants, mostly of the delta variant (Table 2).

**Table 1.**
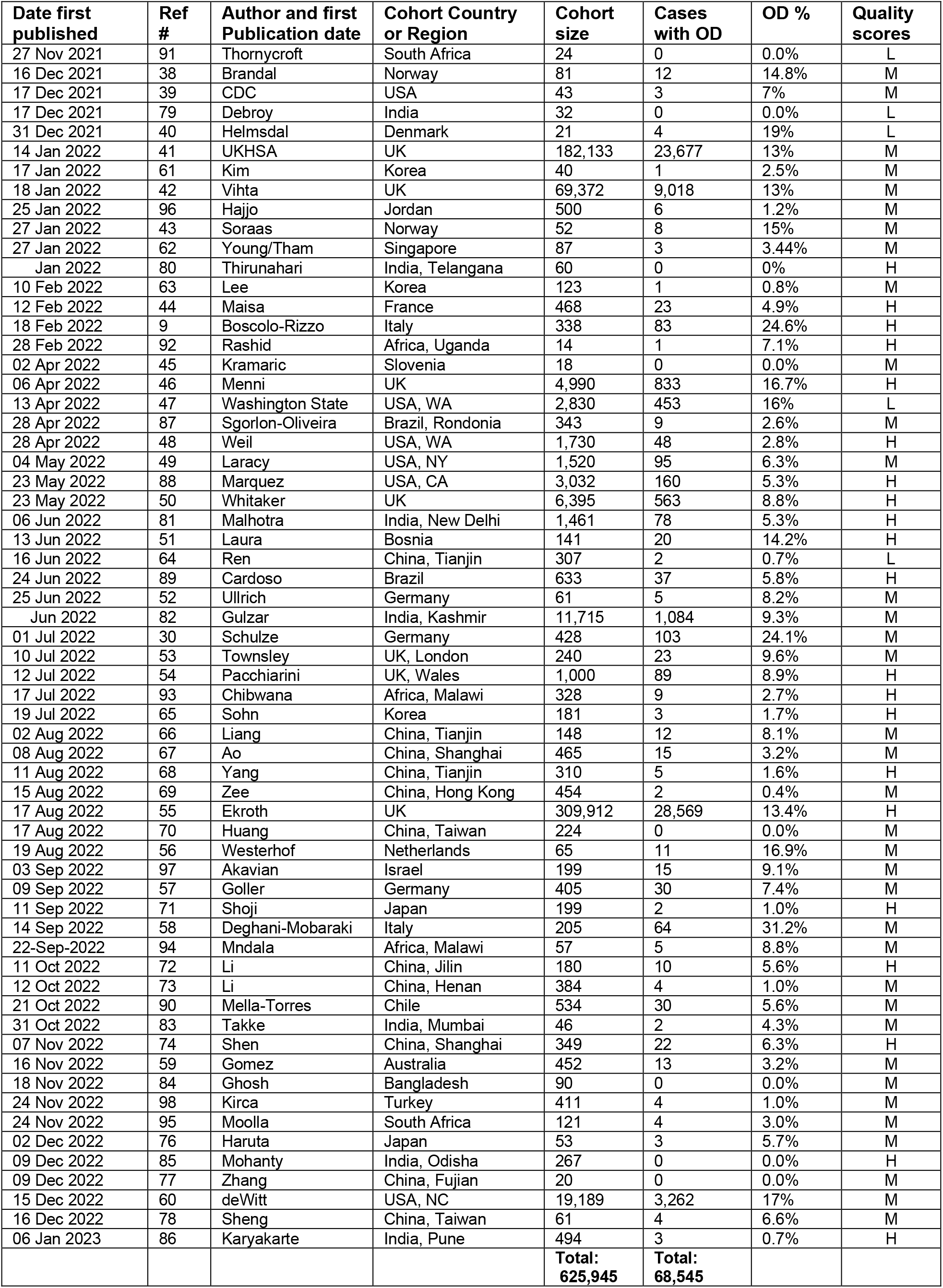
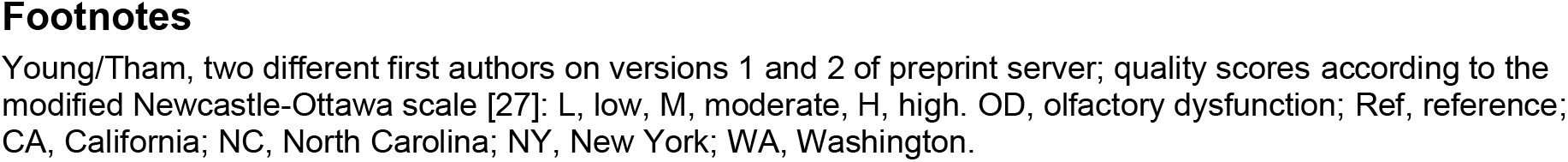
List of studies reporting the prevalence of olfactory dysfunction (OD) caused by the omicron variant.

| Date first published | Ref # | Author and first Publication date | Cohort Country or Region | Cohort size | Cases with OD | OD % | Quality scores |
| --- | --- | --- | --- | --- | --- | --- | --- |
| 27 Nov 2021 | 91 | Thornycroft | South Africa | 24 | 0 | 0.0% | L |
| 16 Dec 2021 | 38 | Brandal | Norway | 81 | 12 | 14.8% | M |
| 17 Dec 2021 | 39 | CDC | USA | 43 | 3 | 7% | M |
| 17 Dec 2021 | 79 | Debroy | India | 32 | 0 | 0.0% | L |
| 31 Dec 2021 | 40 | Helmsdal | Denmark | 21 | 4 | 19% | L |
| 14 Jan 2022 | 41 | UKHSA | UK | 182,133 | 23,677 | 13% | M |
| 17 Jan 2022 | 61 | Kim | Korea | 40 | 1 | 2.5% | M |
| 18 Jan 2022 | 42 | Vihta | UK | 69,372 | 9,018 | 13% | M |
| 25 Jan 2022 | 96 | Hajjo | Jordan | 500 | 6 | 1.2% | M |
| 27 Jan 2022 | 43 | Soraas | Norway | 52 | 8 | 15% | M |
| 27 Jan 2022 | 62 | Young/Tham | Singapore | 87 | 3 | 3.44% | M |
| Jan 2022 | 80 | Thirunahari | India, Telangana | 60 | 0 | 0% | H |
| 10 Feb 2022 | 63 | Lee | Korea | 123 | 1 | 0.8% | M |
| 12 Feb 2022 | 44 | Maisa | France | 468 | 23 | 4.9% | H |
| 18 Feb 2022 | 9 | Boscolo-Rizzo | Italy | 338 | 83 | 24.6% | H |
| 28 Feb 2022 | 92 | Rashid | Africa, Uganda | 14 | 1 | 7.1% | H |
| 02 Apr 2022 | 45 | Kramaric | Slovenia | 18 | 0 | 0.0% | M |
| 06 Apr 2022 | 46 | Menni | UK | 4,990 | 833 | 16.7% | H |
| 13 Apr 2022 | 47 | Washington State | USA, WA | 2,830 | 453 | 16% | L |
| 28 Apr 2022 | 87 | Sgorlon-Oliveira | Brazil, Rondonia | 343 | 9 | 2.6% | M |
| 28 Apr 2022 | 48 | Weil | USA, WA | 1,730 | 48 | 2.8% | H |
| 04 May 2022 | 49 | Laracy | USA, NY | 1,520 | 95 | 6.3% | M |
| 23 May 2022 | 88 | Marquez | USA, CA | 3,032 | 160 | 5.3% | H |
| 23 May 2022 | 50 | Whitaker | UK | 6,395 | 563 | 8.8% | H |
| 06 Jun 2022 | 81 | Malhotra | India, New Delhi | 1,461 | 78 | 5.3% | H |
| 13 Jun 2022 | 51 | Laura | Bosnia | 141 | 20 | 14.2% | H |
| 16 Jun 2022 | 64 | Ren | China, Tianjin | 307 | 2 | 0.7% | L |
| 24 Jun 2022 | 89 | Cardoso | Brazil | 633 | 37 | 5.8% | H |
| 25 Jun 2022 | 52 | Ullrich | Germany | 61 | 5 | 8.2% | M |
| Jun 2022 | 82 | Gulzar | India, Kashmir | 11,715 | 1,084 | 9.3% | M |
| 01 Jul 2022 | 30 | Schulze | Germany | 428 | 103 | 24.1% | M |
| 10 Jul 2022 | 53 | Townsley | UK, London | 240 | 23 | 9.6% | M |
| 12 Jul 2022 | 54 | Pacchiarini | UK, Wales | 1,000 | 89 | 8.9% | H |
| 17 Jul 2022 | 93 | Chibwana | Africa, Malawi | 328 | 9 | 2.7% | H |
| 19 Jul 2022 | 65 | Sohn | Korea | 181 | 3 | 1.7% | H |
| 02 Aug 2022 | 66 | Liang | China, Tianjin | 148 | 12 | 8.1% | M |
| 08 Aug 2022 | 67 | Ao | China, Shanghai | 465 | 15 | 3.2% | M |
| 11 Aug 2022 | 68 | Yang | China, Tianjin | 310 | 5 | 1.6% | H |
| 15 Aug 2022 | 69 | Zee | China, Hong Kong | 454 | 2 | 0.4% | M |
| 17 Aug 2022 | 55 | Ekroth | UK | 309,912 | 28,569 | 13.4% | H |
| 17 Aug 2022 | 70 | Huang | China, Taiwan | 224 | 0 | 0.0% | M |
| 19 Aug 2022 | 56 | Westerhof | Netherlands | 65 | 11 | 16.9% | M |
| 03 Sep 2022 | 97 | Akavian | Israel | 199 | 15 | 9.1% | M |
| 09 Sep 2022 | 57 | Goller | Germany | 405 | 30 | 7.4% | M |
| 11 Sep 2022 | 71 | Shoji | Japan | 199 | 2 | 1.0% | H |
| 14 Sep 2022 | 58 | Deghani-Mobaraki | Italy | 205 | 64 | 31.2% | M |
| 22-Sep-2022 | 94 | Mndala | Africa, Malawi | 57 | 5 | 8.8% | M |
| 11 Oct 2022 | 72 | Li | China, Jilin | 180 | 10 | 5.6% | H |
| 12 Oct 2022 | 73 | Li | China, Henan | 384 | 4 | 1.0% | M |
| 21 Oct 2022 | 90 | Mella-Torres | Chile | 534 | 30 | 5.6% | M |
| 31 Oct 2022 | 83 | Takke | India, Mumbai | 46 | 2 | 4.3% | M |
| 07 Nov 2022 | 74 | Shen | China, Shanghai | 349 | 22 | 6.3% | H |
| 16 Nov 2022 | 59 | Gomez | Australia | 452 | 13 | 3.2% | M |
| 18 Nov 2022 | 84 | Ghosh | Bangladesh | 90 | 0 | 0.0% | M |
| 24 Nov 2022 | 98 | Kirca | Turkey | 411 | 4 | 1.0% | M |
| 24 Nov 2022 | 95 | Moolla | South Africa | 121 | 4 | 3.0% | M |
| 02 Dec 2022 | 76 | Haruta | Japan | 53 | 3 | 5.7% | M |
| 09 Dec 2022 | 85 | Mohanty | India, Odisha | 267 | 0 | 0.0% | H |
| 09 Dec 2022 | 77 | Zhang | China, Fujian | 20 | 0 | 0.0% | M |
| 15 Dec 2022 | 60 | deWitt | USA, NC | 19,189 | 3,262 | 17% | M |
| 16 Dec 2022 | 78 | Sheng | China, Taiwan | 61 | 4 | 6.6% | M |
| 06 Jan 2023 | 86 | Karyakarte | India, Pune | 494 | 3 | 0.7% | H |
|  |  |  |  | **Total: 625,945** | **Total: 68,545** |  |  |

**Table 2.** Compilation of the 32 studies that compare prevalence of olfactory dysfunction due to omicron with that due to delta or other variants.

| Region | Ref # | Author | Country or Region | Cohort Size | Percent hyposmia | Cohort Size | Percent hyposmia | Reduction Om./Prev. | Variant Name |
| --- | --- | --- | --- | --- | --- | --- | --- | --- | --- |
|  |  |  |  | Omicron |  | Previous variants |  |  |  |
| Middle East | 97 | Akavian | Israel | 199 | 9.1% | 119 | 51.3% | 17.5% | G614 |
| Middle East | 98 | Kirca | Turkey | 411 | 1% | 960 | 5.8% | 17.2% | wt |
| Africa | 95 | Moola | South Africa | 121 | 3.3% | 116 | 9.5% | 34.7% | G614 |
| Africa | 93 | Chibwana | Malawi | 328 | 2.7% | 154 | 5.8% | 46.6% | δ ? |
| Africa | 94 | Mndala | Malawi | 57 | 8.8% | 128 | 10.2% | 86.3% | δ |
| East Asia | 62 | Young/Tham | Singapore | 87 | 3.44% | 87 | 2.3% | 149.6% | δ |
| East Asia | 68 | Yang | China, Tianjin | 310 | 1.6% | 422 | 6.9% | 23.2% | β, δ |
| East Asia | 73 | Li | China, Jilin | 384 | 1% | 103 | 2% | 50.0% | δ |
| East Asia | 71 | Shoji | Japan | 199 | 1.0% | 111 | 19% | 5.3% | δ |
| East Asia | 70 | Huang | China, Taiwan | 224 | 0.0% | 141 | 4.3% | 0.0% | α |
| South Asia | 84 | Ghosh | Bangladesh | 90 | 0.0% | 40 | 10.0% | 0.0% | δ |
| South Asia | 85 | Mohanty | India, Odisha | 267 | 0.0% | 461 | 3.2% | 0.0% | δ ? |
| South Asia | 83 | Takke | India, Mumbai | 46 | 0.43% | 55 | 10.9% | 39.4% | δ |
| South Asia | 81 | Malhotra | India, New Delhi | 1,461 | 5.3% | 1,907 | 51.6% | 10.3% | δ |
| Hispanic | 88 | Marquez | USA, CA | 3,032 | 5.3% | 1,533 | 18.2% | 29.1% | δ, prev. |
| Latino | 89 | Cardoso | Brazil | 633 | 5.8% | 5,420 | 48.2% | 12.0% | wt, γ, δ |
| Latino | 90 | Mella-Torres | Chile | 534 | 5.6% | 54 | 13% | 43.1% | δ |
| Western | 41 | UKHSA | UK | 182,133 | 13% | 87,920 | 34% | 38.2% | δ |
| Western | 42 | Vihta | UK | 69,372 | 13% | 14,318 | 40% | 32.5% | δ |
| Western | 43 | Soraas | Norway | 52 | 15% | 18 | 72.2% | 20.8% | δ |
| Western | 9 | Boscolo-Rizzo | Italy | 338 | 24.6% | 441 | 62.6% | 39.3% | G614 |
| Western | 46 | Menni | UK | 4,990 | 16.7% | 4,990 | 52.7% | 31.7% | δ |
| Western | 48 | Weil | USA, WA | 1,730 | 2.8% | 209 | 11.1% | 25.2% | δ |
| Western | 49 | Laracy | USA, NY | 1,520 | 6.3% | 361 | 29% | 21.7% | α, δ |
| Western | 50 | Whitaker | UK | 6,395 | 8.8% | 6,739 | 16.2% | 54.3% | wt, α, δ |
| Western | 30 | Schulze | Germany | 428 | 24.1% | 1,497 | 66.7% | 36.1% | G614, α, δ |
| Western | 54 | Pacciarini | UK, Wales | 1,000 | 8.9% | 8,168 | 25.8% | 34.5% | δ |
| Western | 55 | Ekroth | UK | 309,912 | 13.4% | 123,529 | 33.7% | 39.8% | δ |
| Western | 56 | Westerhof | Netherlands | 65 | 16.9% | 216 | 46.7% | 36.2% | G614, α |
| Western | 59 | Gomez | Australia | 452 | 3.2% | 425 | 36.9% | 8.7% | δ |
| Western | 53 | Townsley | UK, London | 240 | 9.6% | 67 | 37.3% | 25.7% | δ |
| Western | 60 | deWitt | USA, NC | 19,189 | 17% | 37,711 | 55% | 30.9% | pre δ |

**Figure 2.**
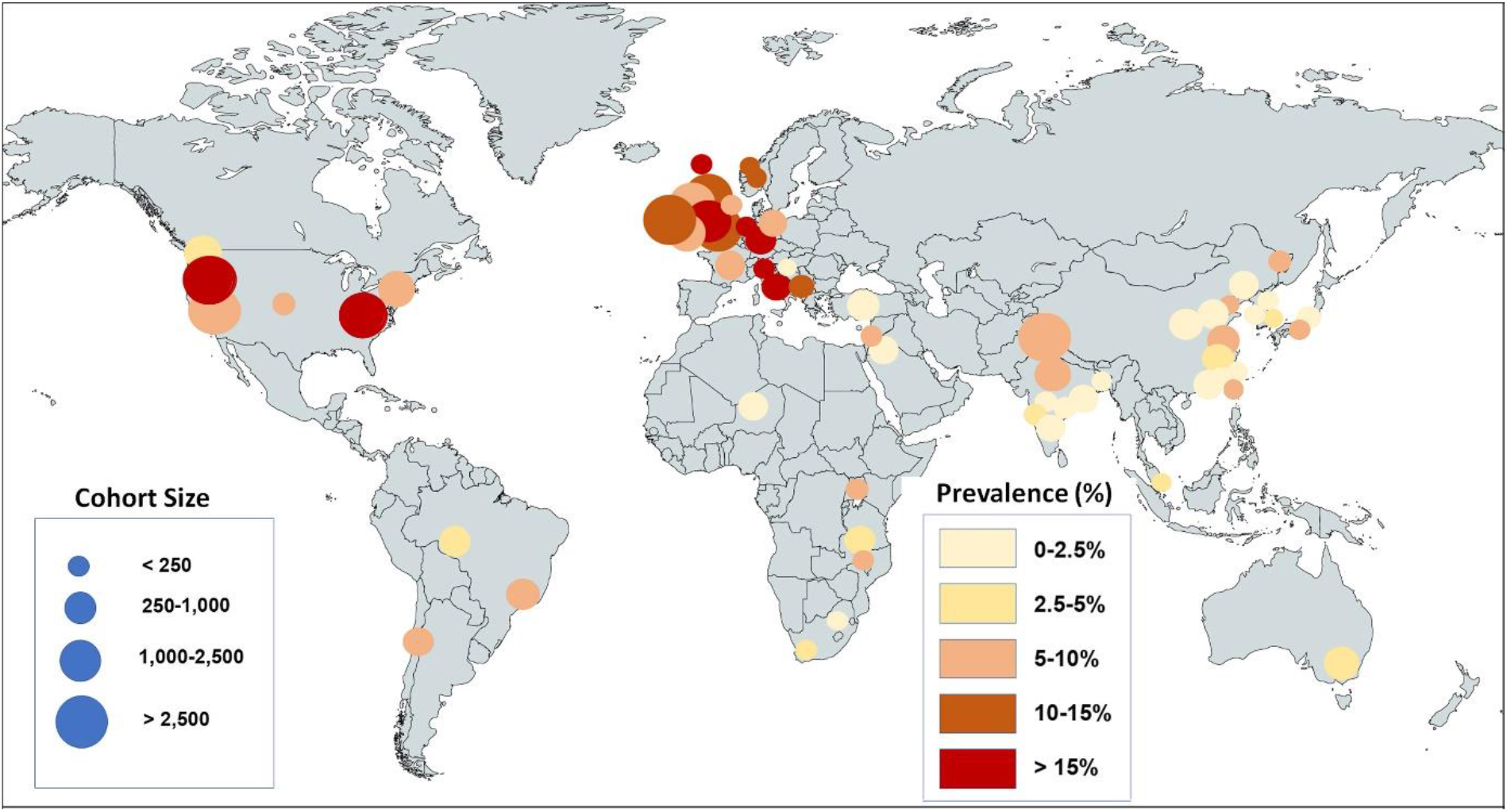
World map showing the location of cohorts included in the systematic review and the prevalence of olfactory dysfunction due to the omicron variant. The size of the circles represents the size of the cohort as indicated in blue; the color gradient indicates the prevalence range as shown on the right side. Note that populations of European ancestry have larger prevalences than populations of non-European ancestry.

### 3.2 Global Prevalence of Olfactory Dysfunction

When we combine all eligible studies in the Forest Plot (Fig. 3), we derive an estimate of the global prevalence of olfactory dysfunction due to the omicron variant as being 6.6% of adults infected with this variant. However, this estimate obscures that ethnicity is a major factor. Our meta-analysis of the studies reporting on populations of European ancestry, which are the majority of studies and the ones with the largest cohort sizes, shows that the pooled prevalence of olfactory dysfunction is 11.7% (Fig. 4). On the other hand, populations of non-European ancestry have a much lower prevalence, ranging from 1.9% to 4.9%, as detailed below (Fig. 4). When ethnic differences between populations and the current population sizes are weighted appropriately (Fig. 4; Table 3), the global prevalence of olfactory dysfunction due to the omicron variant reduces to 3.7% of omicron-infected adults.

**Figure 3.**
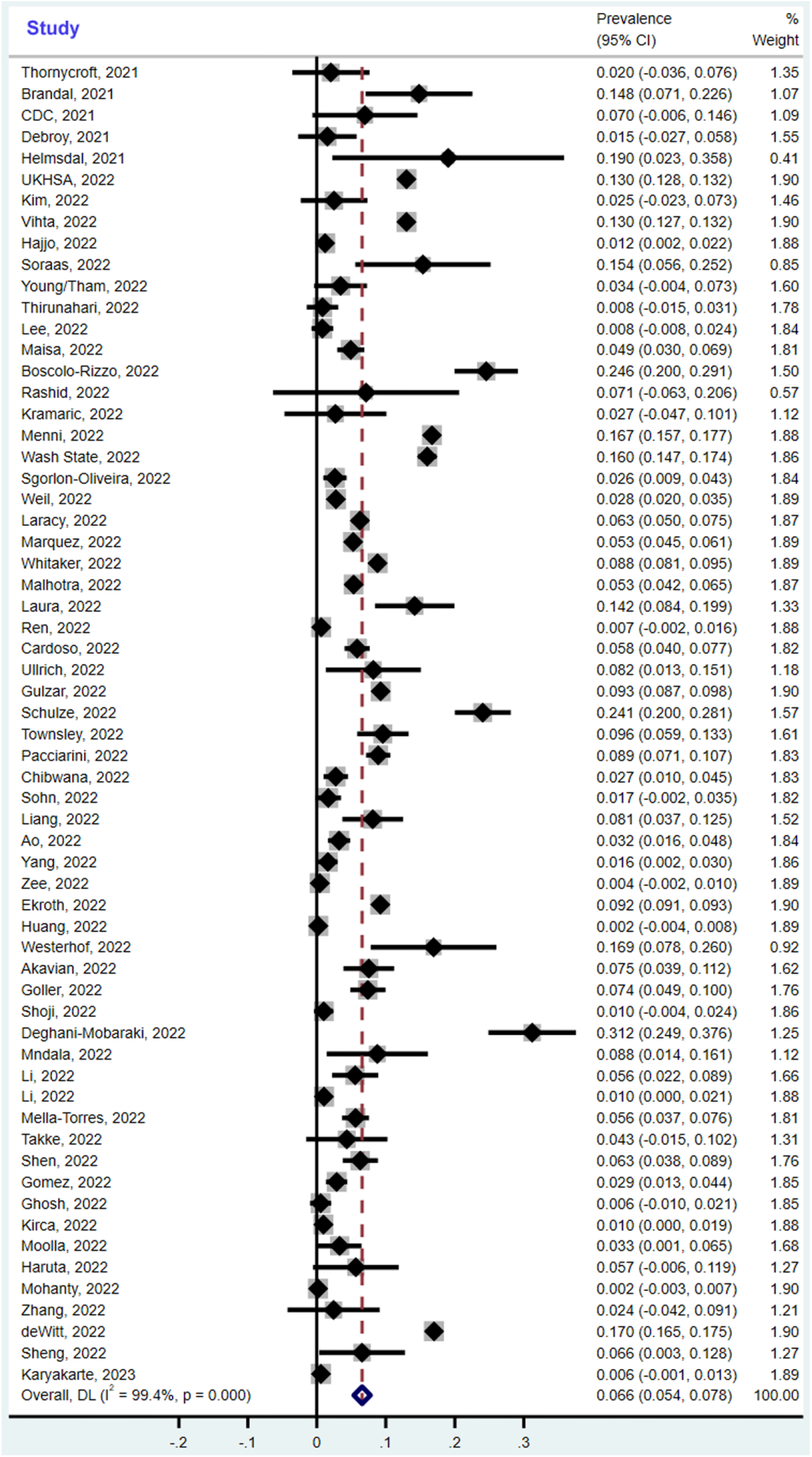
Forest plot of the 62 studies reporting the prevalence of olfactory dysfunction due to the omicron variant. The confidence intervals (CI) and the weight of each study are indicated on the right. The pooled overall global prevalence is 6.6% according to the meta-analysis, but this does not take into account ethnic differences and population sizes as explained in Fig. 4. The size of the light grey box is proportional to the study weight. The study weights are obtained based on the DerSimonian-Laird method. CI, confidence interval; DL, DerSimonian-Laird method; I^2^, I-squared index.

**Figure 4.**
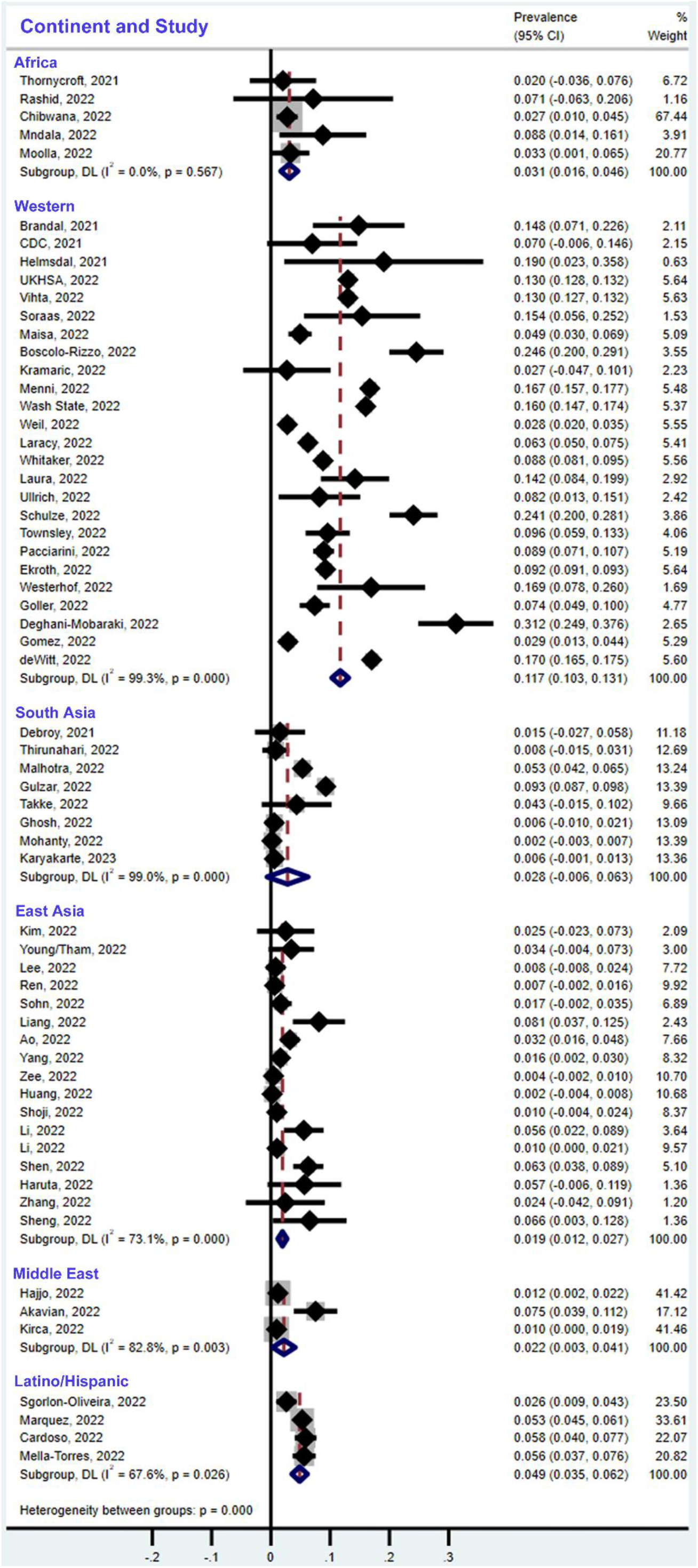
Forest plots of the prevalence of olfactory dysfunction due to omicron differentiated by regions/ethnic populations according to the meta-analysis. Prevalences are 3.1% in populations in Africa (CI = 1.6%-4.6%), 11.7% in people of European ancestry (Western countries, CI = 10.3%-13.1%), 2.8% in South Asia (CI = 0%-6.3%), 1.9% in East Asia (CI = 1.2%-2.7%), 2.2% in the Middle East (CI = 0.3%-4.1%), and 4.9% in Latinos/Hispanics (CI = 3.5%-6.2%). The size of the light grey box is proportional to the study weight. The study weights are obtained based on the DerSimonian-Laird method. CI, confidence interval; DL, DerSimonian-Laird method; I^2^, I-squared index.

**Table 3.** Estimation of the number of adults in different ethnicities expected to experience olfactory dysfunction (OD) with omicron.

|  | Population | Adults only | COVID Infected Adults * | OD Prevalence | Adults with OD | Weight | Prevalence x Weight |
| --- | --- | --- | --- | --- | --- | --- | --- |
|  | billion | billion | billion | % | million |  |  |
| Western | 0.9 | 0.675 | 0.6075 | 11.7 | 71.1 | 0.11 | 1.32 |
| Latino/ Hispanic | 0.7 | 0.525 | 0.4725 | 4.9 | 23.2 | 0.09 | 0.43 |
| Africa | 1.4 | 1.050 | 0.9450 | 3.1 | 29.3 | 0.18 | 0.54 |
| East Asia | 2.5 | 1.875 | 1.6875 | 1.9 | 32.1 | 0.31 | 0.59 |
| South Asia | 2.0 | 1.500 | 1.3500 | 2.8 | 37.8 | 0.25 | 0.70 |
| Middle East | 0.5 | 0.375 | 0.3375 | 2.2 | 7.4 | 0.06 | 0.14 |
| Total | 8.0 | 6.00 | 5.40 |  | 200.9 | 1.00 | 3.72 |

\* COVID-infected = 90% for all populations (IHME, October 21, 2022 [6])
Weight = (number of COVID patients in one continent) / (number of total COVID patients)

When we compared the prevalence of olfactory dysfunction due to omicron with that of previous variants (mostly delta), we find a 2-fold to 10-fold lower prevalence with omicron, based on the 32 studies that provided a direct comparison (Table 2; Fig. 5A, B). The overall reduction of olfactory dysfunction associated with omicron vs. previous variants is 0.299 (confidence intervals (CIs): 0.276, 0.323). This difference is statistically significant (p<0.001). When compared with previous meta-analyses reporting on multiple SARS-CoV-2 variants up to August 15, 2020 (prevalence: 43.0%, 104 studies with 38,198 patients [99]) and up to November 10, 2020 (prevalence: 38.2%, 107 studies with 32,142 patients [100]), the prevalence of olfactory dysfunction due to omicron is 10-fold lower (Fig. 5B). The funnel plots (Fig. 6A, B) indicate that the included studies do not demonstrate publication bias (A, p=0.591; B, p=0.703).

**Figure 5.**
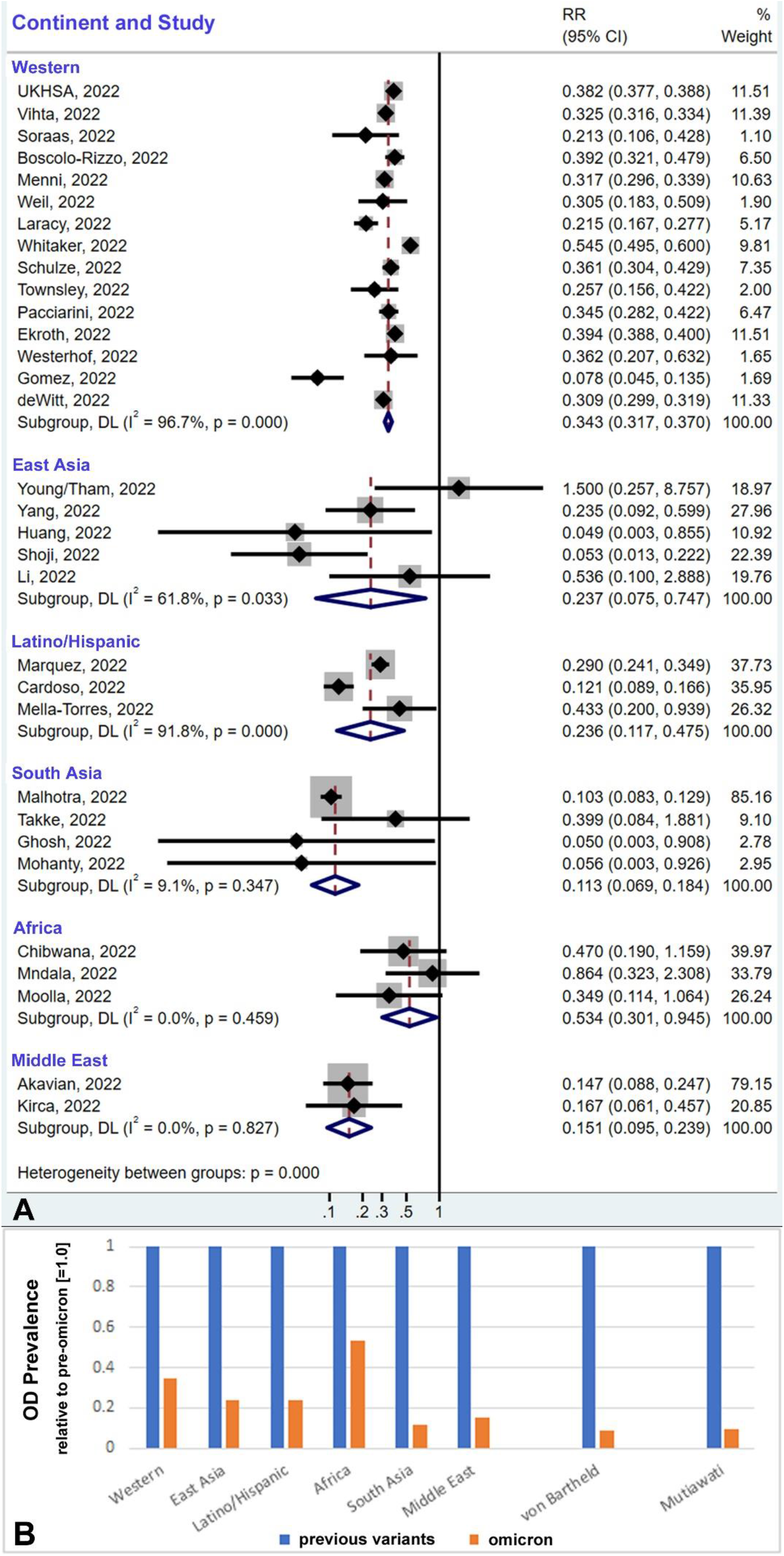
The prevalence of olfactory dysfunction (OD) due to omicron is reduced by 2-fold to 10-fold compared to the previous SARS-CoV-2 variants, varying by ethnic population and region. (A) The reduction of OD due to omicron in direct comparisons within similar populations and regions during the predominance of mostly the delta variant (for specifics of the comparator variants, see Table 2). The size of the light grey box is proportional to the study weight. CI, confidence interval; DL, DerSimonian-Laird method; I^2^, I-squared index, RR, risk ratio. (B) The bar graph summarizes the reduction in prevalence of OD for the direct comparisons from panel A (n=32), and also two indirect comparisons with pooled estimates from previous meta-analyses, von Bartheld et al., 2020 [99], and Mutiawati et al., 2021 [100]. The percent reduction ranges between 2-fold and 10-fold.

**Figure 6.**
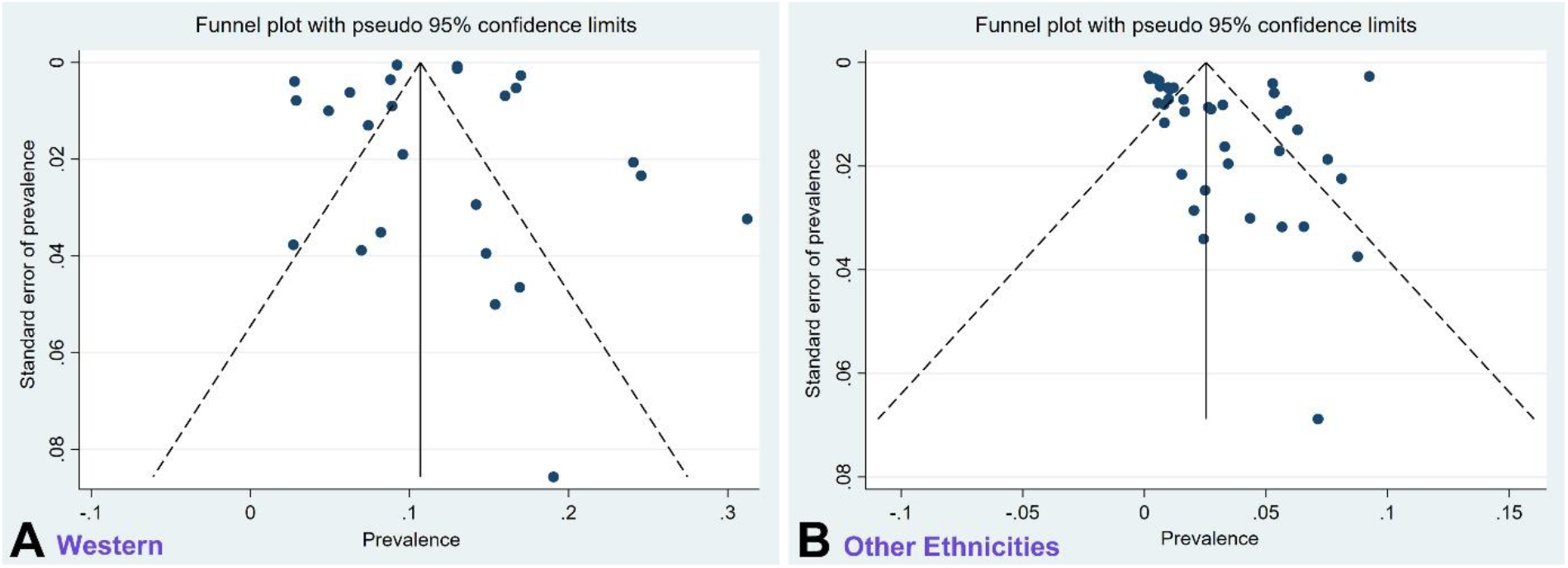
Funnel plots for the Western studies (A) and the studies for all other ethnicities (B) reporting omicron-induced olfactory dysfunction. These are scatterplots of prevalences against their standard errors. The vertical solid line is the estimated effect size; the dotted lines are the corresponding pseudo 95% confidence intervals (CIs). They provide insight into the spread of observed effect sizes. The majority of studies are randomly scattered within the CI region, indicating absence of publication bias for Western studies (A, p=0.591) and for all other ethnicities (B, p=0.703).

### 3.3 Geographic/Ethnic Differences

The studies compiled in Figs. 2 and 4 suggest that geography or ethnicity is a relevant variable. Most ethnicities are well represented, with a robust number of studies and cohort sizes for Western countries (mostly people of European ancestry, n=25 studies, with 602,089 people in all cohorts), for South Asia (n=8 studies, with 14,165 people in all cohorts), East Asia (n=17 studies, with 3,585 people in all cohorts), and for Hispanics/Latinos (n=4 studies, with 4,542 people in all cohorts). The population data are sparse for the African continent (n=5 studies, with 544 people in all cohorts) and in the Middle East (n=3 studies, with 1,100 people in all cohorts). A comparison of the subgroups indicates that omicron causes a hyposmia prevalence of 1.9% in East Asia (CI = 1.2%-2.7%), 3.1% in Africa (CI = 1.6%-4.6%), 2.2% in the Middle East (CI = 0.3%-4.1%), 2.8% in South Asia (CI = 0%-6.3%), 4.9% in Latinos/Hispanics (CI = 3.5%-6.2%), and 11.7% in Western countries (people of European ancestry, CI = 10.3%-13.1%) (Fig. 4). Since these data are derived from people infected with the same virus variant, the population difference must be primarily due to host factors rather than virus factors, as detailed in the Discussion.

### 3.4 Global Prevalence considering Ethnic Differences and Population Sizes

Taking into account the omicron-caused prevalence of hyposmia for the different major ethnicities, and the total population size of these major ethnicities (obtained from the WHO website: https://www.worldometers.info/geography/7-continents/), and using the estimated numbers of COVID cases from the Institute for Health Metrics and Evaluation [6], we can estimate the number of adults in different ethnic populations that can be expected to experience olfactory dysfunction due to omicron infection (Table 3). Since children make up approximately 25% of the world population, we subtracted 25% from each of the population sizes to account for children – which are not included in our review, because there are too few studies reporting on olfactory dysfunction in omicron-infected children, and children with COVID are known to have much less olfactory dysfunction than adults [25]. Assuming a COVID infection of 90% among populations [6], we predict for people of European ancestry a total number of 71.1 million adults with hyposmia out of 0.6 billion (11.7% prevalence), 23.2 million adults with hyposmia out of 0.5 billion Latinos/Hispanics (4.9% prevalence), 29.3 million adults with hyposmia out of 0.9 billion Africans (3.1% prevalence), 32.1 million adults with hyposmia out of 1.7 billion East Asians (1.9% prevalence), 37.8 million adults with hyposmia out of 1.3 billion South Asians (2.8% prevalence), and 7.4 million adults with hyposmia out of 0.3 billion in the Middle East (2.2% prevalence), adding to a total of 200.9 million adult people with olfactory dysfunction, as summarized in Table 3. The estimates for East Asians take into account that the “Zero-COVID” policy in China has ended, meaning that China is expected to have a 90% infection rate as estimated for the rest of the world [6]. The estimated numbers for East Asia would have been substantially lower during the “Zero-COVID” policy in China.

### 3.5 Ethnic Profiles

Omicron’s Hyposmia vs. UGT2A1 Risk Allele Frequency Initially, it was thought that differences in expression levels or in genetic variation of the virus entry proteins, ACE2 and TMPRSS2, may be host factors that contribute to population differences in COVID’s olfactory dysfunction [11,12]. However, this hypothesis appears to be inconsistent with recent data [14], and it is now thought that the host factor most likely is an odorant-metabolizing enzyme, a glycosyltransferase that is encoded by the UGT2A1/A2 locus, based on a recent genome-wide association study showing significant ethnic differences in the frequency of the risk allele at this locus [13].

The ethnicity profiles (East Asian, African, South Asian, Latino/Hispanic, and people with European ancestry) for both, the risk allele frequency as well as the hyposmia prevalence, is shown in Fig. 7. Our comparison of the major ethnicities for omicron’s hyposmia prevalence reveals a remarkably similar ethnic profile when compared with the pattern described [13] for the frequency of the risk allele in the UGT2A1 locus (Fig. 7). We used meta regression to test whether the risk allele in the UGT2A1 locus predicted omicron’s hyposmia prevalence and we found that there is an association between a population’s risk allele frequency and omicron’s hyposmia prevalence (p<0.001). The coefficient is positive, which means that the hyposmia prevalence is higher when the risk allele frequency is higher, consistent with the genome-wide association analysis [13]. This supports the idea that the odorant-metabolizing enzyme, the UDP glycosyltransferase, is involved as a host factor in the susceptibility to SARS-CoV-2 induced olfactory dysfunction.

**Figure 7.**
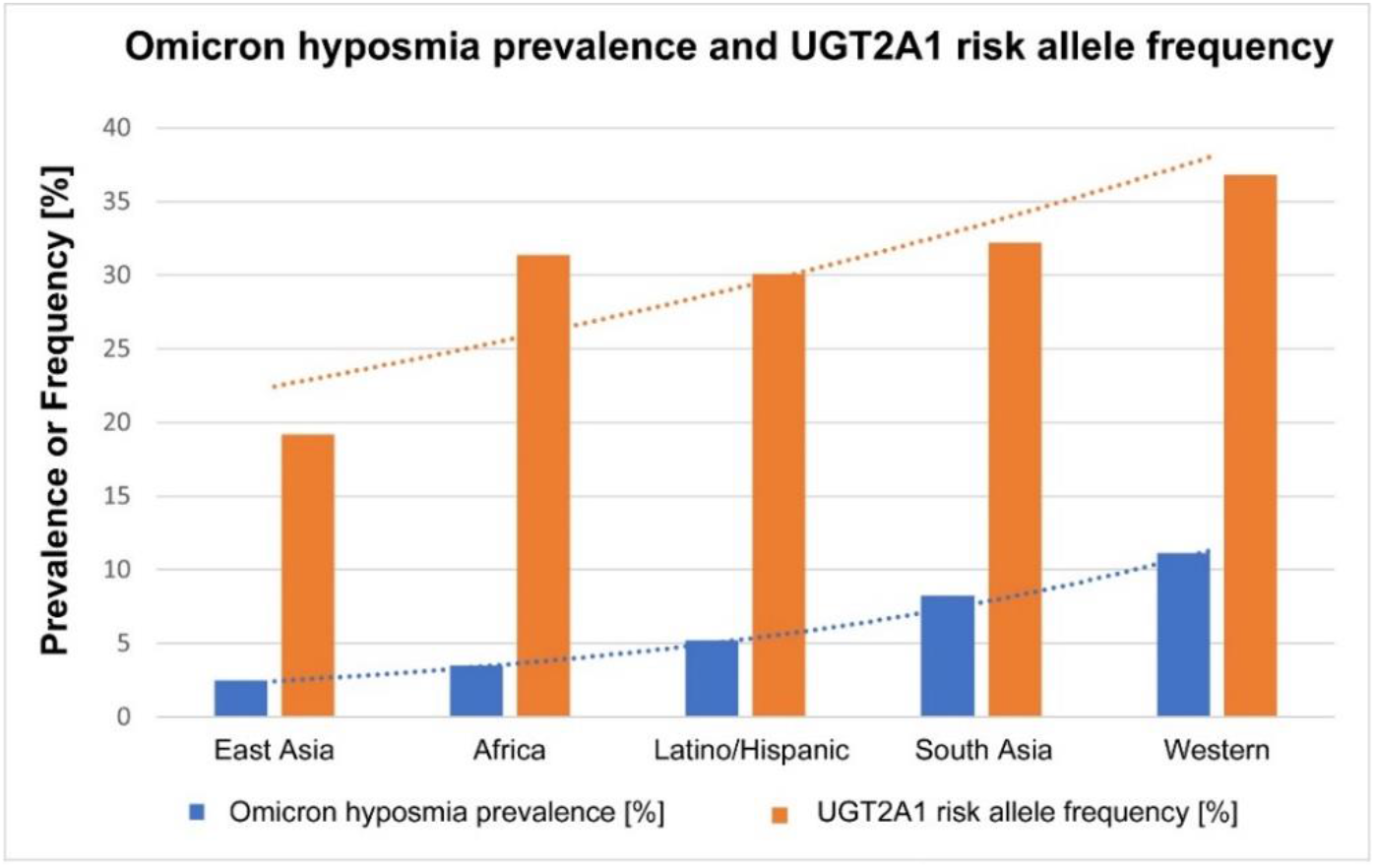
The bar graph shows the differences between ethnicities for omicron’s prevalence of olfactory dysfunction (blue bars), compared with the frequency of the risk allele for olfactory dysfunction in the UGT2A1 locus according to Shelton et al., 2022 [13] (orange bars). The blue bars show the pooled estimate for the hyposmia prevalence of the same ethnicities according to our systematic review. Meta regression shows that there is a positive association between the two parameters (p<0.001). The prevalence of olfactory dysfunction is higher in those ethnic populations that have a higher frequency of the risk allele. The two exponential trend lines show this similarity.

### 3.6 Comparison of survey-type studies and traditional-design studies

It has been cautioned that survey-type studies (that invite people to respond to questionnaires, often posted on the internet) may have a bias, because people with more severe conditions tend to be more motivated to respond [9,28-30,36]. Therefore, we estimated the magnitude of such a potential bias by comparing survey-type studies with traditional-design studies, for people with European ancestry, considering both omicron-caused hyposmia as well as hyposmia caused by previous SARS-CoV-2 variants (Fig. 8A, B). We find that, with omicron, the survey-based studies result in a pooled estimate of the hyposmia prevalence of 14.2% (CI: 9.7%-18.7%), which is higher than the 10.9% (CI: 9.3%-12.6%) with traditional studies (Fig. 8A). However, a meta regression analysis shows that there is no statistically significant difference between these two prevalences (p=0.391). With the previous variants, mostly delta, the survey-based studies result in a pooled estimate of the hyposmia prevalence of 45.4% (CI: 22.1%-68.8%), which is higher than the 36.6% (CI: 28.4%-44.8%) with traditional studies (Fig. 8B). A meta regression analysis again shows that there is no statistically significant difference between these two prevalences (p=0.428).

**Figure 8.**
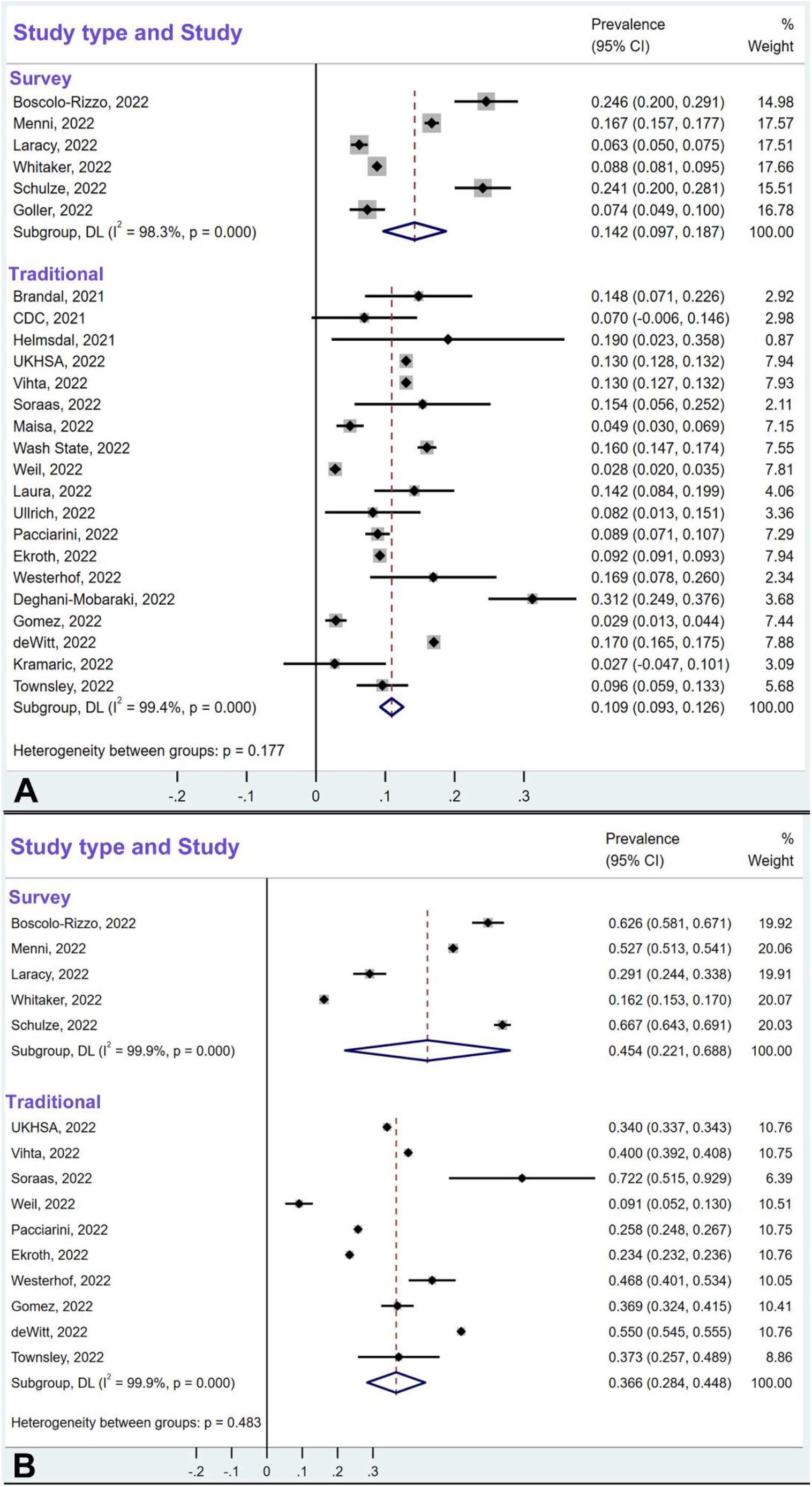
The pooled prevalences of olfactory dysfunction due to omicron (A) and due to other variants (B) in survey-type studies and traditional-design studies for populations of European ancestry. While the pooled estimates are higher in the survey-type studies than in the traditional-design studies,14.2% vs.10.9% for omicron studies (A), and 45.4% vs 36.6% for previous variants (B), meta-regression showed no heterogeneity between the two study types. Confidence intervals for omicron studies (A) were 9.7%-18.7% for survey-type studies, and 9.3%-12.6% for traditional design studies. Confidence intervals for previous variant studies (B) were 22.1%-68.8% for survey-type studies, and 28.4%-44.8% for traditional design studies.

## 4. Discussion

### 4.1 Global Prevalence of Olfactory Dysfunction with Omicron

We estimate the global prevalence of omicron-induced olfactory dysfunction in adults to be 3.7%. This estimate takes into account ethnic differences and population sizes and is based on the notion that 90% of the population has been or will be exposed to SARS-CoV-2 [6]. Our estimate of 3.7% prevalence translates into 200.9 million adults predicted to experience omicron-induced olfactory dysfunction (Table 3). Our review and meta-analysis show that the olfactory dysfunction after omicron infection is 2-10-fold lower than with previous variants, with a substantial reduction in all ethnicities (Fig. 5B).

Our analysis reveals significant ethnic differences in the prevalence of omicron-induced olfactory dysfunction: The estimation of omicron’s current olfactory dysfunction in Western countries with 11.7% prevalence is well supported, and the estimates for South Asians, Latinos/Hispanics, and East Asians, with 14,165, 4,542, and 3,585 people in the cohorts, respectively, are also fairly well attested. Our estimates about the prevalence in populations in Africa (3.1%) and the Middle East (2.2%) are less certain, as they are based on only five and three studies, respectively, with 544 and 1,100 people in the cohorts.

### 4.2 Why is Omicron’s Effect on Olfaction Different than that of Previous Variants?

Two main reasons have been proposed to explain this phenomenon, and they are not mutually exclusive. The mutations in the spike protein make the omicron variant more hydrophobic [101] which may reduce the solubility of the virus in the mucus, diminishing its ability to reach the olfactory epithelium [2,102]. Second, due to reduced furin cleavage, the omicron variant prefers an endosomal route via cathepsin for entering host cells rather than a surface membrane fusion via the protease TMPRSS2 [103]. Sustentacular cells and Bowman gland cells are the cells in the olfactory epithelium which most abundantly express not only ACE2, but also TMPRSS2 [104,105] and for this reason these support cells were the prime target of previous SARS-CoV-2 variants for host cell entry via the route using cell surface membrane fusion enabled by TMPRSS2 [106]. Since the support cells – similar to many other host cells – have evolved more potent defense mechanisms for the endosomal route of infection [102,107], for example, the antiviral *IFITM2* gene is the most highly upregulated gene in support cells at 3 days after infection [108], this may lead to a lower efficiency in omicron infection of the support cells of the olfactory epithelium, and therefore reduced olfactory dysfunction [102,109].

### 4.3 Ethnic Differences in UGT2A1 Risk Allele Frequency: Implications

The similarity in the ethnic profiles between omicron’s prevalence of hyposmia and the frequency of the UGT2A1 risk allele (Fig. 7) suggests that the UGT2A1-encoded glycosyltransferase is the host factor, or one of the major host factors, that determines the risk of olfactory dysfunction due to SARS-CoV-2 infection. How does the UDP glycosyltransferase affect the sense of smell? This is an evolutionary highly conserved enzyme in olfaction, not only from rodents to human [16,18], but also in invertebrates [15]. It is thought to modulate the concentration of odorant molecules and terminate odorant signal transduction [17,110]. It contributes to the biotransformation of odorant molecules, prevents saturation of the odorant receptors, modifies the perceived quality of odorants [16,110], and thereby plays a major role in olfactory sensitivity. Polymorphisms in the enzyme may account for inter-individual variability in olfactory perception [16]. Furthermore, UDP glycosyltransferases are expressed differentially with aging [110] which could explain the increased olfactory dysfunction seen in young adults (but less in children or older people [25,99]), and expression of UDP glycosyltransferase also differs between genders [111], which may explain the higher female susceptibility to olfactory dysfunction [99].

What does the genetic/ethnic difference in the risk allele frequency in the host (with the most extreme values in East Asians vs European ancestry) tell us about olfactory dysfunction? The risk allele at the UGT2A1 locus causes more olfactory dysfunction [13], which may explain why Europeans are more susceptible to loss of smell, but the mechanism still is unclear. Nevertheless, the new data point to the sustentacular cell as the site of pathogenesis, directing us towards a better understanding of how SARS-CoV-2 attacks the olfactory system.

### 4.4 Technical Considerations of Methodology

Similar to pre-omicron COVID research where the large majority of studies reported olfactory dysfunction based on patient recall [27], we also had to rely on studies reporting the results of such subjective testing (patient recall) for the monitoring of olfactory dysfunction. In fact, we found only a single study that reported omicron-induced hyposmia based on psychophysical testing [66], while there are 62 studies that report such dysfunction based on subjective recall (Table 1). It is currently controversial whether psychophysical testing or subjective recall is the most valid and sensitive approach to assess COVID-related chemosensory dysfunction [21,27]. Among studies that compared the two types of methods, about half of them concluded that objective testing is more sensitive than subjective testing [112-115]. The other half concluded the opposite, that subjective testing is more sensitive than objective testing [116-120]. While some authors recommend psychophysical testing as being superior to subjective patient recall [121,122], others have pointed out that for the assessment of olfactory dysfunction in the COVID pandemic, psychophysical testing by ENT specialists is largely impractical, because people with an acute COVID infection typically quarantine during the acute phase. Furthermore, since chemosensory loss often lasts only about a week [99,123], smell and taste may have recovered before they can be tested quantitatively by experts [21]. Another argument against psychophysical testing is that its interpretation requires not only cross-cultural validation [22,124], but also a pre-pandemic or pre-infection base level for each individual, because of the large fraction (nearly 30%) of people with pre-existing olfactory dysfunction in the normal population when measured by this method [125]. However, such a pre-pandemic base level is rarely available.

The large majority of the studies we compiled were scored as moderate or high quality according to the modified Newcastle-Ottawa scale [27] (Table 1), and omission of studies scoring low for quality did not change our results and conclusions. There was no evidence for publication bias in the analysis of the funnel plots (Fig. 6A, B). Because people who are more impacted by their condition may be more likely to respond in an internet-based survey [9,28-30,36], this could lead to bias in survey-type studies. Therefore, we compared survey-type studies with traditional representative sampling studies that use direct and immediate questioning of each member of the eligible cohort, rather than inviting eligible individuals online and collecting responses on internet-provided questionnaires. Such study designs rely on equitable participation of individuals suffering from loss of smell and those with no such loss. We found that although survey-type studies reported higher prevalence of hyposmia than traditional study designs, there was no evidence for heterogeneity between the two study types (Fig. 8A, B). The issue of potential bias in survey-type studies deserves further scrutiny and should be examined in the future with more studies and larger cohorts.

### 4.5 Limitations of our Review

Most studies compiled in our review did not stratify by age group, but age is a relevant factor [13,99,126]. Likewise, most studies did not report on gender of the cohort and gender of the cases, yet gender is also a relevant factor [13,99]. There were few studies from Africa and the Middle East, and those studies had small cohorts – additional data are needed to conduct a more reliable subgroup analysis and achieve higher certainty for the prevalence of hyposmia in these populations.

Many studies did not specify change in smell vs change in taste, and reported them as either change or loss of smell *and* taste, or change or loss of smell *or* taste. Additional studies are needed to better distinguish effects of omicron on smell and taste. We did not attempt to resolve whether omicron sub-variants have different effects on olfactory dysfunction – there are too few studies yet that report effects of subvariants on loss of smell [48,50,57,86].

Some cohorts of the studies were ethnically mixed, but the exact ethnic composition of the cohort was reported in only a few studies (e.g., [48]), and none of the studies reported the prevalence of olfactory dysfunction separately for distinct ethnicities. This should be done in the future to verify differences between ethnicities, and this may also “sharpen” the ethnic distinctions which may be blurred by ethnically mixed cohorts. For example, the large fraction of Asians and/or Latinos/Hispanics in the cohorts of Weil et al. [48] and (likely) of Laracy et al. [49] may explain the relatively low overall prevalence of hyposmia in their studies. Although ethnic patterns are emerging, more detailed analyses in future studies may allow to assign a more precise prevalence of olfactory dysfunction to each major ethnicity.

We did not include studies that focused on olfactory dysfunction in children. Children have a lower hyposmia prevalence than adults [25], and the reduction of the prevalence with omicron would be more difficult to quantify, given the considerable ethnic differences. Nevertheless, to our knowledge, there are six studies that reported children’s prevalence of olfactory dysfunction with omicron [127-132], and all of these studies except for one [130] found a substantial reduction (about five-fold) of olfactory dysfunction with omicron when compared with previous variants which is similar to the relative sparing of olfaction by omicron in adults (Fig. 5).

### 4.6 Future Directions

Although we have a clue that the UGT2A1 locus, and therefore the UDP glycosyltransferase, is involved in the ethnic differences in COVID-related olfactory dysfunction [13], the mechanism still is unclear. Nevertheless, the sustentacular support cell in the olfactory epithelium appears to play a major role. This helps to direct focus on this cell type and its key roles in the processing of odorants and fundamental workings of the sense of smell [102]. A better understanding of the molecular mechanisms of loss of smell in COVID may inform about new therapies to help with persistent loss of smell, beyond the current olfactory training that is not effective for more than half of cases [133].

While some studies indicate that a previous SARS-CoV-2 infection may reduce the likelihood of olfactory dysfunction in a subsequent COVID infection [51], it is known that a previous COVID infection with an earlier variant does not necessarily prevent a second loss of smell when the same individual subsequently becomes infected with a different SARS-CoV-2 variant [134,135]. It also does not seem that vaccinations reliably prevent the occurrence of loss of smell in break-through infections [9,52,88,136].

If omicron infects about 90% of 6 billion adult people worldwide, what does a global prevalence of 3.7% olfactory dysfunction (200.9 million cases world-wide) mean for trends in global cases of olfactory dysfunction? Our ethnicity-adjusted projections suggest that olfactory dysfunction will decline globally despite the higher infectivity of the omicron variant, contrary to previous predictions [9,10]. It is not yet known whether there will be *persistent* loss of smell after omicron infection. Will it be similar to previous variants – with about 5% persistent loss of smell among those who experience olfactory dysfunction [10]? Does a lower number of cases of olfactory dysfunction with omicron also reduce the percentage of those who will have a persistent loss of smell? We don’t know yet about persistent loss of smell caused by omicron, since it has been only a little more than one year since the first cases of omicron infection emerged. Much is still to be learned about the effects of omicron (and previous and future) variants of SARS-CoV-2 on olfaction.

## Data Availability

All data produced in the present work are contained in the manuscript.

## Acknowledgments

We thank the following colleagues for helpful discussions: Hans Vasquez-Gross and Juli Petereit (Nevada Bioinformatics Center, RRID:SCR_017802, University of Nevada, Reno), Adam Auton (23andMe, Sunnyvale, California), and Nicolas Meunier (University of Saclay, Paris).

## Author Contributions

Both authors contributed to the data collection, analysis, and writing of the manuscript.

## Data Availability Statement

All data referenced in this study are publicly available.

## Graphical Abstract

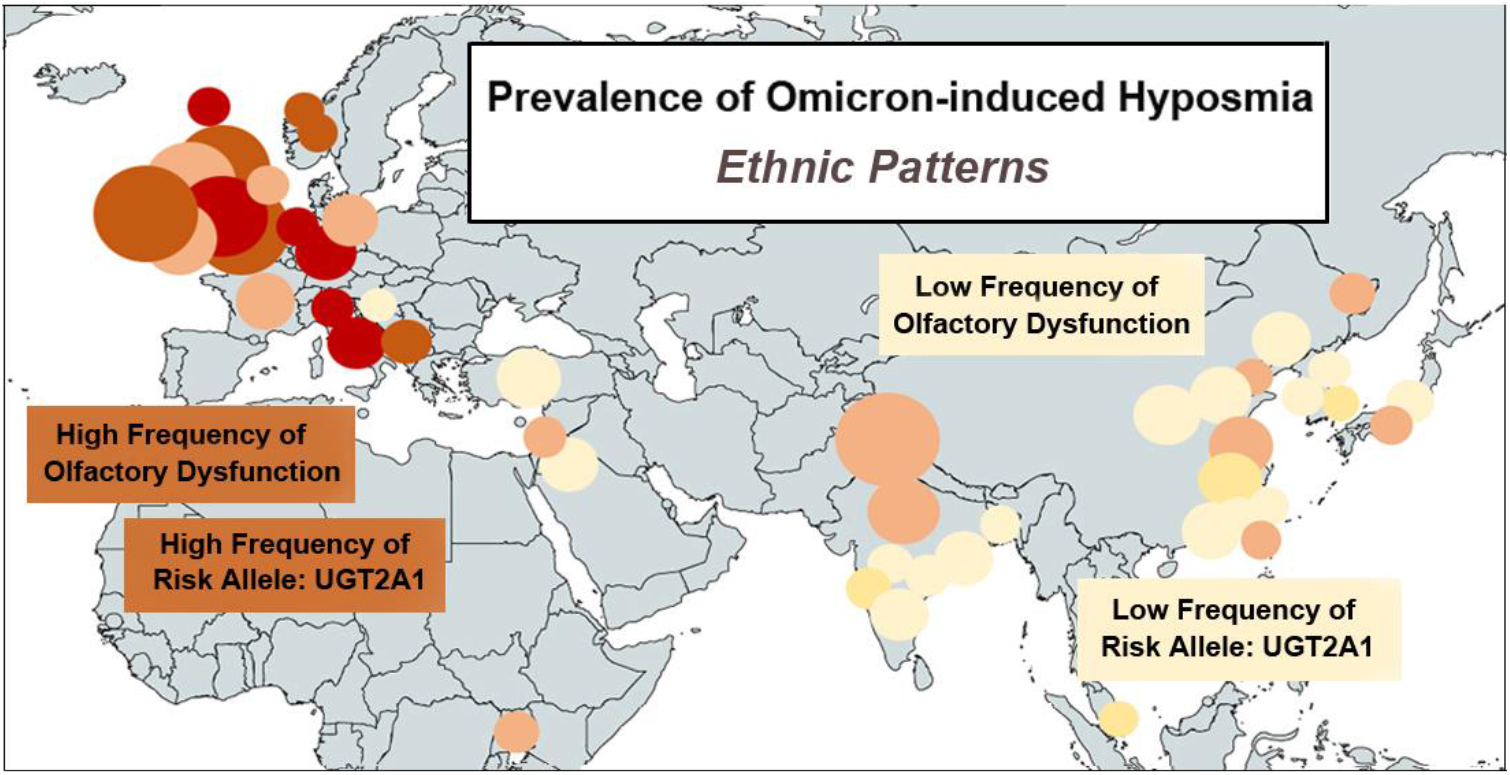

## References

1. Rodriguez-Sevilla, J.J.; Güerri-Fernádez, R.; Bertran Recasens, B. Is There Less Alteration of Smell Sensation in Patients with Omicron SARS-CoV-2 Variant Infection? Front. Med. (Lausanne). 2022, 9, 852998. doi: 10.3389/fmed.2022.852998.

2. Butowt, R.; Bilińska, K.; von Bartheld, C. Why Does the Omicron Variant Largely Spare Olfactory Function? Implications for the Pathogenesis of Anosmia in Coronavirus Disease 2019. J. Infect. Dis. 2022, 226(8):1304-1308. doi: 10.1093/infdis/jiac113.

3. Patt, Y.S.; David, P.; Bergwerk, M.; Shoenfeld, Y. The Reduced Frequency of Olfactory Dysfunction in Patients with Omicron SARS-CoV-2 Variant Infection. Ann. Otolaryngol. Rhinol. 2022, 9, 1302.

4. Chee, J.; Chern, B.; Loh, W.S.; Mullol, J.; Wang, Y. Pathophysiology of SARS-CoV-2 Infection of Nasal Respiratory and Olfactory Epithelia and Its Clinical Impact. Curr. Allergy Asthma Rep. 2023 Jan 4:1–11. doi: 10.1007/s11882-022-01059-6. Epub ahead of print.

5. COVID-19 Cumulative Infection Collaborators. Estimating global, regional, and national daily and cumulative infections with SARS-CoV-2 through Nov 14, 2021: a statistical analysis. Lancet 2022, 399, 2351–2380. doi: 10.1016/S0140-6736(22)00484-6.

6. IHME (Institute for Health Metrics and Evaluation). COVID-19 Results Briefing, Global, October 21, 2022. https://www.healthdata.org/sites/default/files/files/Projects/COVID/2022/1_briefing_Global_10.pdf accessed on December 13, 2022

7. von Bartheld, C.S.; Hagen, M.M.; Butowt, R. The D614G Virus Mutation Enhances Anosmia in COVID-19 Patients: Evidence from a Systematic Review and Meta-analysis of Studies from South Asia. ACS Chem. Neurosci. 2021, 12, 3535–3549. doi: 10.1021/acschemneuro.1c00542.

8. See, A.; Ko, K.K.K.; Toh, S.T. Epidemiological analysis in support of hypothesis that D614G virus mutation is a major contributing factor to chemosensory dysfunction in COVID-19 patients. Eur. Arch. Otorhinolaryngol. 2021, 278, 3595–3596. doi: 10.1007/s00405-021-06941-z.

9. Boscolo-Rizzo, P.; Tirelli, G.; Meloni, P.; Hopkins, C.; Madeddu, G.; De Vito, A.; Gardenal, N.; Valentinotti, R.; Tofanelli, M.; Borsetto, D., et al. Coronavirus disease 2019 (COVID-19)-related smell and taste impairment with widespread diffusion of severe acute respiratory syndrome-coronavirus-2 (SARS-CoV-2) Omicron variant. Int. Forum Allergy Rhinol. 2022, 12, 1273–1281. doi: 10.1002/alr.22995.

10. Tan, B.K.J.; Han, R.; Zhao, J.J.; Tan, N.K.W.; Quah, E.S.H.; Tan, C.J.; Chan, Y.H.; Teo, N.W.Y.; Charn, T.C.; See, A.; et al. Prognosis and persistence of smell and taste dysfunction in patients with covid-19: meta-analysis with parametric cure modelling of recovery curves. BMJ 2022, 378, e069503. doi: 10.1136/bmj-2021-069503.

11. Butowt, R., Bilinska, K.; von Bartheld, C.S. Chemosensory Dysfunction in COVID-19: Integration of Genetic and Epidemiological Data Points to D614G Spike Protein Variant as a Contributing Factor. ACS Chem. Neurosci. 2020, 11, 3180–3184. doi: 10.1021/acschemneuro.0c00596

12. Kumar, A.A.; Lee, S.W.Y.; Lock, C.; Keong, N.C. Geographical Variations in Host Predisposition to COVID-19 Related Anosmia, Ageusia, and Neurological Syndromes. Front. Med. (Lausanne). 2021, 8, 661359. doi: 10.3389/fmed.2021.661359.

13. Shelton, J.F.; Shastri, A.J.; Fletez-Brant, K.; 23andMe COVID-19 Team; Aslibekyan, S., Auton, A. The UGT2A1/UGT2A2 locus is associated with COVID-19-related loss of smell or taste. Nat. Genet. 2022, 54, 121–124. doi: 10.1038/s41588-021-00986-w.

14. Braga-Paz, I.; Ferreira de Araújo, J.L.; Alves, H.J.; de Ávila, R.E.; Resende, G.G.; Teixeira, M.M.; de Aguiar, R.S.; de Souza, R.P.; Bahia, D. Negative correlation between ACE2 gene expression levels and loss of taste in a cohort of COVID-19 hospitalized patients: New clues to long-term cognitive disorders. Front. Cell. Infect. Microbiol. 2022, 12, 905757. doi: 10.3389/fcimb.2022.905757.

15. Heydel, J.; Leclerc, S.; Bernard, P.; Pelczar, H.; Gradinaru, D.; Magdalou, J.; Minn, A.; Artur, Y.; Goudonnet, H. Rat olfactory bulb and epithelium UDP-glucuronosyltransferase 2A1 (UGT2A1) expression: in situ mRNA localization and quantitative analysis. Brain Res. Mol. Brain Res. 2001, 90, 83–92. doi: 10.1016/s0169-328x(01)00080-8.

16. Heydel, J.M.; Coelho, A.; Thiebaud, N.; Legendre, A.; Le Bon, A.M.; Faure, P.; Neiers, F., Artur, Y.; Golebiowski, J.; Briand, L. Odorant-binding proteins and xenobiotic metabolizing enzymes: implications in olfactory perireceptor events. Anat. Rec. (Hoboken) 2013, 296, 1333–45. doi: 10.1002/ar.22735.

17. Lazard, D.; Zupko, K.; Poria, Y.; Nef, P.; Lazarovits, J.; Horn, S.; Khen, M.; Lancet, D. Odorant signal termination by olfactory UDP glucuronosyl transferase. Nature 1991, 349, 790–793. doi: 10.1038/349790a0.

18. Schwartz, M.; Menetrier, F.; Heydel, J.M.; Chavanne, E.; Faure, P.; Labrousse, M.; Lirussi, F.; Canon, F.; Mannervik, B.; Briand, L.; Neiers, F. Interactions Between Odorants and Glutathione Transferases in the Human Olfactory Cleft. Chem. Senses 2020, 45, 645–654. doi: 10.1093/chemse/bjaa055.

19. Khan, M.; Yoo, S.J.; Clijsters, M.; Backaert, W.; Vanstapel, A.; Speleman, K.; Lietaer, C.; Choi, S.; Hether, T.D.; Marcelis, L.; et al. Visualizing in deceased COVID-19 patients how SARS-CoV-2 attacks the respiratory and olfactory mucosae but spares the olfactory bulb. Cell 2021, 184, 5932-5949.e15. doi: 10.1016/j.cell.2021.10.027.

20. Finlay, J.B.; Brann, D.H.; Abi-Hachem, R.; Jang, D.W.; Oliva, A.D.; Ko, T.; Gupta, R.; Wellford, S.A.; Moseman, E.A.; Jang, S.S.; et al. Persistent post-COVID-19 smell loss is associated with immune cell infiltration and altered gene expression in olfactory epithelium. Sci. Transl. Med. 2022, 14, eadd0484. doi: 10.1126/scitranslmed.add0484.

21. Hintschich, C.A.; Niv, M.Y.; Hummel, T. The taste of the pandemic-contemporary review on the current state of research on gustation in coronavirus disease 2019 (COVID-19). Int. Forum Allergy Rhinol. 2022, 12, 210–216. doi: 10.1002/alr.22902.

22. Langstaff, L.; Clark, A.; Salam, M.; Philpott, C.M. Cultural Adaptation and Validity of the Sniffin’ Sticks Psychophysical Test for the UK Setting. Chem. Percept. 2021,14, 102–108. https://doi.org/10.1007/s12078-021-09287-2

23. Moher, D.; Liberati, A.; Tetzlaff, J.; Altman, D.G; PRISMA Group. Preferred reporting items for systematic reviews and meta-analyses: the PRISMA statement. BMJ. 2009, 339, b2535. doi: 10.1136/bmj.b2535.

24. Hansen, C.; Perofsky, A.C.; Burstein, R.; Famulare, M.; Boyle, S.; Prentice, R.; Marshall, C.; McCormick, B.J.J.; Reinhart, D.; Capodanno, B.; et al. Trends in Risk Factors and Symptoms Associated with SARS-CoV-2 and Rhinovirus Test Positivity in King County, Washington, June 2020 to July 2022. JAMA Netw. Open. 2022, 5, e2245861. doi: 10.1001/jamanetworkopen.2022.45861

25. Yadav, M.; Singh, A.; Meena, J.; Sankar, J.M. A systematic review and meta-analysis of otorhinolaryngological manifestations of coronavirus disease 2019 in paediatric patients. J. Laryngol. Otol. 2022, 136, 588–603. doi: 10.1017/S0022215122000536.

26. Coelho, D.H.; Reiter, E.R.; French, E.; Costanzo, R.M. Decreasing Incidence of Chemosensory Changes by COVID-19 Variant. Otolaryngol. Head Neck Surg. 2022 May 3:1945998221097656. doi: 10.1177/01945998221097656.

27. Boscutti, A.; Delvecchio, G.; Pigoni, A.; Cereda, G.; Ciappolino, V.; Bellani, M.; Fusar-Poli, P.; Brambilla, P. Olfactory and gustatory dysfunctions in SARS-CoV-2 infection: A systematic review. Brain Behav. Immun. Health 2021, 15, 100268. doi: 10.1016/j.bbih.2021.100268.

28. Lerner, D.K.; Garvey, K.L.; Arrighi-Allisan, A.E.; Filimonov, A.; Filip, P.; Shah, J.; Tweel, B.; Del Signore, A.; Schaberg, M.; Colley, P.; et al. Clinical Features of Parosmia Associated with COVID-19 Infection. Laryngoscope 2022, 132, 633–639. doi: 10.1002/lary.29982.

29. Ohla, K.; Veldhuizen, M.G.; Green, T.; Hannum, M.E.; Bakke, A.J.; Moein, S.T.; Tognetti, A.; Postma, E.M.; Pellegrino, R.; Hwang, D.L.D.; et al. A follow-up on quantitative and qualitative olfactory dysfunction and other symptoms in patients recovering from COVID-19 smell loss. Rhinology 2022 Apr 10. doi: 10.4193/Rhin21.415. Epub ahead of print.

30. Schulze, H.; Bayer, W. Changes in Symptoms Experienced by SARS-CoV-2-Infected Individuals – From the First Wave to the Omicron Variant. Front. Virol. 2022, 2, 880707. doi: 10.3389/fviro.2022.880707

31. Egger, M.; Davey Smith, G.; Schneider, M.; Minder, C. Bias in meta-analysis detected by a simple, graphical test. BMJ 1997, 315, 629–34. doi: 10.1136/bmj.315.7109.629.

32. Korber, B.; Fischer, W.M.; Gnanakaran, S.; Yoon, H.; Theiler, J.; Abfalterer, W.; Hengartner, N.; Giorgi, E.E.; Bhattacharya, T.; Foley, B.; et al. Tracking Changes in SARS-CoV-2 Spike: Evidence that D614G Increases Infectivity of the COVID-19 Virus. Cell 2020, 182, 812-827.e19. doi: 10.1016/j.cell.2020.06.043.

33. Higgins, J.P.; Thompson, S.G. Quantifying heterogeneity in a meta-analysis. Stat. Med. 2002, 21, 1539–1558.

34. DerSimonian, R.; Laird, N. Meta-analysis in clinical trials. Control Clin. Trials. 1986, 7, 177–188.

35. Higgins, J.P.T. Cochrane Collaboration. Cochrane handbook for systematic reviews of interventions. Second edition. ed. Hoboken, NJ: Wiley-Blackwell; 2020.

36. Hood, K.; Robling, M.; Ingledew, D.; Gillespie, D.; Greene, G.; Ivins, R.; Russell, I.; Sayers, A.; Shaw, C.; Williams, J. Mode of data elicitation, acquisition and response to surveys: a systematic review. Health Technol. Assess. 2012, 16, 1–162. doi: 10.3310/hta16270.

37. Egger, M.; Smith, G.D.; Phillips, A.N. Meta-analysis: principles and procedures. BMJ. 1997, 315, 1533–1537.

38. Brandal, L.T.; MacDonald, E.; Veneti, L.; Ravlo, T.; Lange, H.; Naseer, U.; Feruglio, S.; Bragstad, K.; Hungnes, O.; Ødeskaug, L.E.; et al. Outbreak caused by the SARS-CoV-2 Omicron variant in Norway, November to December 2021. Euro. Surveill. 2021, 26, 2101147. doi: 10.2807/1560-7917.ES.2021.26.50.2101147.

39. CDC COVID-19 Response Team. SARS-CoV-2 B.1.1.529 (Omicron) Variant - United States, December 1-8, 2021. MMWR Morb. Mortal. Wkly. Rep. 2021 70, 1731–1734. doi: 10.15585/mmwr.mm7050e1.

40. Helmsdal, G.; Hansen, O.K.; Møller, L.F.; Christiansen, D.H.; Petersen, M.S.; Kristiansen, M.F. Omicron Outbreak at a Private Gathering in the Faroe Islands, Infecting 21 of 33 Triple-Vaccinated Healthcare Workers. Clin. Infect. Dis. 2022, 75, 893–896. doi: 10.1093/cid/ciac089.

41. UK Health Security Agency. SARS-CoV-2 variants of concern and variants under investigation in England. Technical briefing 34. 2022. https://assets.publishing.service.gov.uk/government/uploads/system/uploads/attachment_data/file/1046853/technical-briefing-34-14-january-2022.pdf xAccessed December 13, 2022

42. Vihta, K.D.; Pouwels, K.B.; Peto, T.E.; Pritchard, E.; House, T.; Studley, R.; Rourke, E.; Cook, D.; Diamond, I.; Crook, D.; et al. Omicron-associated changes in SARS-CoV-2 symptoms in the United Kingdom. Clin. Infect. Dis. 2022 Aug 3:ciac613. doi: 10.1093/cid/ciac613.

43. Søraas, A.; Grødeland, G.; Granerud, B.K..; Ueland, T.; Lind, A.; Fevang, B.; Murphy, S.L.; Huse, C.; Nygaard, A.B.; Steffensen, A.K.; et al. Breakthrough infections with the omicron and delta variants of SARS-CoV-2 result in similar re-activation of vaccine-induced immunity. Front. Immunol. 2022, 13, 964525. doi: 10.3389/fimmu.2022.964525.

44. Maisa, A.; Spaccaferri, G.; Fournier, L.; Schaeffer, J.; Deniau, J.; Rolland, P.; Coignard, B.; regional COVID-19 investigation team; EMERGEN consortium. First cases of Omicron in France are exhibiting mild symptoms, November 2021-January 2022. Infect. Dis. Now. 2022, 52, 160–164. doi: 10.1016/j.idnow.2022.02.003.

45. Kramarič, J.; Ješe, R.; Tomšič, M.; Rotar, Ž.; Hočevar, A. COVID-19 among patients with giant cell arteritis: a single-centre observational study from Slovenia. Clin. Rheumatol. 2022, 41, 2449–2456. doi: 10.1007/s10067-022-06157-4.

46. Menni, C.; Valdes, A.M.; Polidori, L.; Antonelli, M.; Penamakuri, S.; Nogal, A.; Louca, P.; May, A.; Figueiredo, J.C.; Hu, C.; et al. Symptom prevalence, duration, and risk of hospital admission in individuals infected with SARS-CoV-2 during periods of omicron and delta variant dominance: a prospective observational study from the ZOE COVID Study. Lancet 2022, 399, 1618–1624. doi: 10.1016/S0140-6736(22)00327-0.

47. Washington State Department of Health. SARS-CoV-2 Sequencing and Variants in Washington State April 13, 2022. https://doh.wa.gov/sites/default/files/2022-02/420-316-SequencingAndVariantsReport.pdf accessed May 8, 2022

48. Weil, A.A.; Luiten, K.G.; Casto, A.M.; Bennett, J.C.; O’Hanlon, J.; Han, P.D.; Gamboa, L.S.; McDermot, E.; Truong, M.; Gottlieb, G.S.; et al. Genomic surveillance of SARS-CoV-2 Omicron variants on a university campus. Nat. Commun. 2022; 13, 5240. doi:10.1038/s41467-022-32786-z

49. Laracy, J.C.; Robilotti, E.V.; Yan, J.; Lucca, A.; Aslam, A.; Babady, N.E.; Kamboj, M. Comparison of coronavirus disease 2019 (COVID-19) symptoms at diagnosis among healthcare personnel before and after the emergence of the omicron variant. Infect. Control Hosp. Epidemiol. 2022 May 4:1–3. doi: 10.1017/ice.2022.105. Epub ahead of print.

50. Whitaker, M.; Elliott, J.; Bodinier, B.; Barclay, W.; Ward, H.; Cooke, G.; Donnelly, C.A.; Chadeau-Hyam, M.; Elliott, P. Variant-specific symptoms of COVID-19 in a study of 1,542,510 adults in England. Nat. Commun. 2022, 13, 6856. doi: 10.1038/s41467-022-34244-2.

51. Laura, L.; Dalmatin-Dragišić, M.; Martinović, K.; Tutiš, B.; Herceg, I.; Arapović, M.; Arapović, J. Does pre-existing immunity determine the course of SARS-CoV-2 infection in health-care workers? Single-center experience. Infection 2022, 13, 1–8. doi: 10.1007/s15010-022-01859-y.

52. Ullrich, F.; Hanoun, C.; Turki, A.T.; Liebregts, T.; Breuckmann, K.; Alashkar, F.; Reinhardt, H.C.; von Tresckow, B.; von Tresckow, J. Early report on the severity of COVID-19 in hematologic patients infected with the SARS-CoV2 omicron variant. Eur. J. Haematol. 2022, 109, 364–372. doi: 10.1111/ejh.13818.

53. Townsley, .H.; Carr, E.C.; Russell, T.W.; Adams, L.; Mears, H.V.; Bailey, C.; Black, J.R.M.; Fowler, A.S.; Wilkinson, K.; Hutchinson, M.; et al. Non-hospitalised, vaccinated adults with COVID-19 caused by Omicron BA.1 and BA.2 present with changing symptom profiles compared to those with Delta despite similar viral kinetics. [Preprint] MedRxiv, posted Jul 10, 2022. doi: https://doi.org/10.1101/2022.07.07.22277367

54. Pacchiarini, N.; Sawyer, C.; Williams, C.; Sutton, D.; Roberts, C.; Simkin, F.; King, G.; McClure, V.; Cottrell, S.; Clayton, H.; et al. Epidemiological analysis of the first 1000 cases of SARS-CoV-2 lineage BA.1 (B.1.1.529, Omicron) compared with co-circulating Delta in Wales, UK. Influenza Other Respir. Viruses 2022, 16, 986–993. doi: 10.1111/irv.13021.

55. Ekroth, A.K.E.; Patrzylas, P.; Turner, C.; Hughes, G.J.; Anderson, C. Comparative symptomatology of infection with SARS-CoV-2 variants Omicron (B.1.1.529) and Delta (B.1.617.2) from routine contact tracing data in England. Epidemiol. Infect. 2022, 150, e162. doi: 10.1017/S0950268822001297.

56. Westerhof, I.; de Hoog, M.; Ieven, M.; Lammens, C.; van Beek, J.; Rozhnova, G.; Eggink, D.; Euser, S.; Wildenbeest, J.; Duijts, L.; et al. The impact of variant and vaccination on SARS-CoV-2 symptomatology; three prospective household cohorts. Int. J. Infect. Dis. 2022, 128, 140–147. doi: 10.1016/j.ijid.2022.12.018.

57. Goller, K.V.; Moritz, J.; Ziemann, J.; Kohler, C.; Becker, K.; Hübner, N.O.; The CoMV-Gen Study Group. Differences in Clinical Presentations of Omicron Infections with the Lineages BA.2 and BA.5 in Mecklenburg-Western Pomerania, Germany, between April and July 2022. Viruses 2022, 14, 2033. doi: 10.3390/v14092033

58. Dehgani-Mobaraki, P.; Patel, Z.; Zaidi, A.K.; Giannandrea, D.; Hopkins, C. The Omicron variant of SARS-CoV-2 and its effect on the olfactory system Int. Forum Allergy Rhinol. 2022, [published online ahead of print] 10.1002/alr.23089. doi: 10.1002/alr.23089

59. Gomez, A.; Kelly, M.; Sloan-Gardner, T.S.; Voo, T.V.; Kirk, M.D. Severity and Symptom Characteristics between Omicron and Delta SARS-CoV-2 Variant Infections in the Australian Capital Territory: A Cross-Sectional Study. Research Square [Preprint] Nov 16, 2022, DOI: https://doi.org/10.21203/rs.3.rs-1786210/v1

60. DeWitt, M.E.; Tjaden, A.H.; Herrington, D.; Schieffelin, J.; Gibbs, M.; Weintraub, W.S.; Sanders, J.W.; Edelstein, S.L.; COVID-19 Community Research Partnership. COVID-19 Symptoms by Variant Period in the North Carolina COVID-19 Community Research Partnership, North Carolina, USA. Emerg. Infect. Dis. 2023, 29, 207–211. doi: 10.3201/eid2901.221111.

61. Kim, M.K.; Lee, B.; Choi, Y.Y.; Um, J.; Lee, K.S.; Sung, H.K.; Kim, Y.; Park, J.S.; Lee, M.; Jang, H.C.; et al. Clinical Characteristics of 40 Patients Infected with the SARS-CoV-2 Omicron Variant in Korea. J. Korean Med. Sci. 2022, 37, e31.

62. Tham, S.M.; Fong, S.-W.; Chang, Z.-W.; Tan, K.S.; Rouers, A.; Goh, Y.S.; Tay, D.J.W.; Ong, S.W.X.; Hao, Y.; Chua, S.L., et al. Comparison of the clinical features, viral shedding and immune response in vaccine breakthrough infection by the Omicron and Delta variants. SSRN [Preprint] 2022. Posted on Jun 24, 2022. https://ssrn.com/abstract=4142078

63. Lee, H.Y.; Lee, J.J.; Park, H.; Yu, M.; Kim, J.M.; Lee, S.-E.; Park, Y.-J.; Kim. M.; Kim, S.; Yoo, H.; et al. Importation and community transmission of SARS-CoV-2 B.1.1.529 (Omicron) variant of concern, the Republic of Korea, December 2021. Public Health Weekly Report (PHWR) 2022, 14, 338–343

64. Ren, Y.; Shi, L.; Xie, Y.; Wang, C.; Zhang, W.; Wang, F.; Sun, H.; Huang, L.; Wu, Y.; Xing, Z.; et al. Course and clinical severity of the SARS-CoV-2 Omicron variant infection in Tianjin, China. [Preprint] MedRxiv, posted Jun 16, 2022. doi: https://doi.org/10.1101/2022.06.16.22271932

65. Sohn, Y.J.; Shin, P.J.; Oh, W.S.; Kim, E.; Kim, Y.; Kim, Y.K. Clinical Characteristics of Patients Who Contracted the SARS-CoV-2 Omicron Variant from an Outbreak in a Single Hospital. Yonsei Med. J. 2022, 63, 790–793. doi: 10.3349/ymj.2022.63.8.790.

66. Liang, Y.; Mao, X.; Kuang, M.; Zhi, J.; Zhang, Z.; Bo, M.; Zhang, G.; Lin, P.; Wang, W.; Shen, Z. Interleukin-6 affects the severity of olfactory disorder: a cross-sectional survey of 148 patients who recovered from Omicron infection using the Sniffin’ Sticks test in Tianjin, China. Int. J. Infect. Dis. 2022, 123, 17–24. doi: 10.1016/j.ijid.2022.07.074.

67. Ao, Y.; Li, J.; Wei, Z.; Wang, Z.; Tian, H.; Qiu, Y.; Fu, X.; Ma, W.; Li, L.; Zeng, M.; et al. Clinical and virological characteristics of SARS-CoV-2 Omicron BA.2.2 variant outbreaks during April to May, 2022, Shanghai, China. J. Infect. 2022, 85, 573–607. doi: 10.1016/j.jinf.2022.07.027.

68. Yang, W.; Yang, S.; Wang, L.; Zhou, Y.; Xin, Y.; Li, H.; Mu, W.; Wu, Q.; Xu, L.; Zhao, M.; et al. Clinical characteristics of 310 SARS-CoV-2 Omicron variant patients and comparison with Delta and Beta variant patients in China. Virol. Sin. 2022, 37, 704–715. doi: 10.1016/j.virs.2022.07.014.

69. Zee, S.T.; Kwok, L.F.; Kee, K.M.; Fung, L.H.; Luk, W.P.; Chan, T.L.; Leung, C.P.; Yu, P.W.; Hung, J.; SzeTo, K.Y.; et al. Impact of COVID-19 Vaccination on Healthcare Worker Infection Rate and Outcome during SARS-CoV-2 Omicron Variant Outbreak in Hong Kong. Vaccines (Basel) 2022, 10, 1322. doi: 10.3390/vaccines10081322.

70. Huang, R.C.; Chiu, C.H.; Shang, H.S.; Perng, C.L.; Chiang, T.T.; Tsai, C.C.; Wang, C.H. Clinical characteristics analysis of COVID-19 patients from the first significant community outbreak by SARS-CoV-2 variant B.1.1.7 in Taiwan as experienced from a single northern medical center. J. Microbiol. Immunol. Infect. 2022, 55, 1036–1043. doi: 10.1016/j.jmii.2022.08.007.

71. Shoji, K.; Tsuzuki, S.; Akiyama, T.; Matsunaga, N.; Asai, Y.; Suzuki, S.; Iwamoto, N.; Funaki, T.; Yamada, M.; Ozawa, N.; et al. Comparison of clinical characteristics of COVID-19 in pregnant women between the Delta and Omicron variants of concern predominant periods. J. Infect. Chemother. 2023, 29, 33–38. doi: 10.1016/j.jiac.2022.09.005.

72. Li, H.; Zhu, M.; Zhang, P.; Yan, X.; Niu, J.; Wang, Z.; Cao, J. Milder symptoms and shorter course in patients with re-positive COVID-19: A cohort of 180 patients from Northeast China. Front. Microbiol. 2022, 13, 989879. doi: 10.3389/fmicb.2022.989879.

73. Li, Q.; Liu, X.; Li, L.; Hu, X.; Cui, G.; Sun, R.; Zhang, D.; Li, J.; Li, Y.; Zhang, Y.; et al. Comparison of clinical characteristics between SARS-CoV-2 Omicron variant and Delta variant infections in China. Front. Med. (Lausanne) 2022, 9, 944909. doi: 10.3389/fmed.2022.944909.

74. Shen, J.; Wu, L.; Wang, P.; Shen, X.; Jiang, Y.; Liu, J.; Chen, W. Clinical characteristics and short-term recovery of hyposmia in hospitalized non-severe COVID-19 patients with Omicron variant in Shanghai, China. Front. Med. (Lausanne) 2022, 9, 1038938. doi: 10.3389/fmed.2022.1038938.

75. Shen, X.; Wang, P.; Shen, J.; Jiang, Y.; Wu, L.; Nie, X.; Liu, J.; Chen, W. Neurological Manifestations of hospitalized patients with mild to moderate infection with SARS-CoV-2 Omicron variant in Shanghai, China. J. Infect. Public Health 2022, 16, 155–162. doi: 10.1016/j.jiph.2022.12.005.

76. Haruta, M.; Otsubo, S.; Otsubo, Y. Characteristics of the 6th Japanese wave of COVID-19 in hemodialysis patients. Ren. Replace. Ther. 2022, 8, 61. doi: 10.1186/s41100-022-00451-2.

77. Zhang, H.; Chen, W.; Ye, X.; Zhou, Y.; Zheng, Y.; Weng, Z.; Xie, J.; Zheng, K.; Su, Z.; Zhuang, X.; et al. Clinical characteristics of patients infected with novel coronavirus wild strain, Delta variant strain and Omicron variant strain in Quanzhou: A real world study. Exp. Ther. Med. 2022, 25, 62. doi: 10.3892/etm.2022.11761.

78. Sheng, W.H.; Chang, H.C.; Chang, S.Y.; Hsieh, M.J.; Chen, Y.C.; Wu, Y.Y.; Pan, S.C.; Wang, J.T.; Chen, Y.C. SARS-CoV-2 infection among healthcare workers whom already received booster vaccination during epidemic outbreak of omicron variant in Taiwan. J. Formos. Med. Assoc. 2022 Dec 16:S0929-6646(22)00442-9. doi: 10.1016/j.jfma.2022.12.003.

79. Debroy, S. Omicron-hit had sore throat, body ache, didn’t lose smell. Times of India, December 17, 2021. https://timesofindia.indiatimes.com/city/mumbai/mumbai-omicron-hit-had-sore-throat-body-ache-didnt-lose-smell/articleshow/88327372.cms xAccessed 30 November, 2022

80. Thirunahari, P.S.; Moluguri, A.; Rajamouli, J.; Patruni, M.; Gurnule, S.R. Alarming symptoms in COVID-19 omicron variant, Karimnagar, Telangana. Revista Geintec 2022, 12, 73–80. https://revistageintec.net/wp-content/uploads/2022/09/REVISTA-GEINTEC-6001.pdf

81. Malhotra, S.; Mani, K.; Lodha, R.; Bakhshi, S.; Mathur, V.P.; Gupta, P.; Kedia, S.; Sankar, M.J.; Kumar, P.; Kumar, A.H.V.; et al. COVID-19 infection, and reinfection, and vaccine effectiveness against symptomatic infection among health care workers in the setting of omicron variant transmission in New Delhi, India. Lancet Reg. Health Southeast Asia 2022 Aug;3:100023. doi: 10.1016/j.lansea.2022.100023.

82. Gulzar, B; Rishi, S; Farhana, A; Sheikh, A.A; Dewani, S. Clinico-Demographic characteristics of Third Wave by Omicron (B.1.1.529) variant of SARS-CoV-2 at a Tertiary Care Center, J&K India. J. Adv. Med. Dent. Sci. Res. (Amritsar) 2022 10, 5–9. DOI:10.21276/jamdsr

83. Takke, A.; Zarekar, M.; Muthuraman, V.; Ashar, A.; Patil, K.; Badhavkar, A.; Trivedi, J.; Khargekar, N.; Madkaikar, M.; Banerjee, A. Comparative study of clinical features and vaccination status in Omicron and non-Omicron infected patients during the third wave in Mumbai, India. J. Family Med. Prim. Care. 2022, 11, 6135–6142. doi: 10.4103/jfmpc.jfmpc_430_22.

84. Ghosh, A.K.; Landt, O.; Yeasmin, M.; Sharif, M.; Ratul, R.H.; Molla, M.A.; Nafisa, T.; Mosaddeque, M.B.; Hosen, N.; Bulbul, M.R.H.; et al. Clinical Presentation of COVID-19 and Antibody Responses in Bangladeshi Patients Infected with the Delta or Omicron Variants of SARS-CoV-2. Vaccines (Basel) 2022, 10, 1959. doi: 10.3390/vaccines10111959.

85. Mohanty, M.; Mishra, B.; Singh, A.K.; Mohapatra, P.R.; Gupta, K.; Patro, B.K.; Sahu, D.P.; Kar, P.; Purushotham, P.; Saha, S.; et al. Comparison of Clinical Presentation and Vaccine Effectiveness Among Omicron and Non-omicron SARS Coronavirus-2 Patients. Cureus 2022, 14, e32354. doi: 10.7759/cureus.32354.

86. Karyakarte, R.; Das, R.; Dudhate, S.; Agarasen, J.; Pillai, P.; Chandankhede, P.; Labshetwar, R.; Gadiyal, Y.; Rajmane, M.; Kulkarni, P., et al. Clinical Characteristics and Outcomes of Laboratory-Confirmed SARS-CoV-2 Cases Infected with Omicron subvariants and XBB recombinant variant. [Preprint] MedRxiv posted Jan 6 2023. doi: https://doi.org/10.1101/2023.01.05.23284211

87. Sgorlon-Oliveira, G.; Queiroz, J.A.D.S.; Gasparelo, N.W.F.; Roca, T.P.; Passos-Silva, A.M.; Teixeira, K.S.; Oliveira, A.A.D.S.; Souza, P.R.F.D.; Silva, E.D.S.; Silva, C.C.D.; et al. The SARS-CoV-2 Omicron Variant of Concern and its Rapid Spread throughout the Western Brazilian Amazon. Preprints 2022, 2022040266 (doi: 10.20944/preprints202204.0266.v1). posted 28 Apr 2022

88. Marquez, C.; Kerkhoff, A.D.; Schrom, J.; Rojas, S.; Black, D.; Mitchell, A.; Wang, C.Y.; Pilarowski, G.; Ribeiro, S.; Jones, D.; et al. COVID-19 Symptoms and Duration of Rapid Antigen Test Positivity at a Community Testing and Surveillance Site During Pre-Delta, Delta, and Omicron BA.1 Periods. JAMA Netw. Open 2022, 5, e2235844. doi: 10.1001/jamanetworkopen.2022.35844.

89. Cardoso, C.C.; Rossi, Á.D.; Galliez, R.M.; Faffe, D.S.; Tanuri, A.; Castiñeiras, T.M.P.P. Olfactory Dysfunction in Patients with Mild COVID-19 During Gamma, Delta, and Omicron Waves in Rio de Janeiro, Brazil. JAMA 2022, 328, 582–583. doi: 10.1001/jama.2022.11006.

90. Mella-Torres, A.; Escobar, A.; Barrera-Avalos, C.; Vargas-Salas, S.; Pirazzoli, M.; Gonzalez, U.; Valdes, D.; Rojas, P.; Luraschi, R.; Vallejos-Vidal, E.; et al. Epidemiological characteristics of Omicron and Delta SARS-CoV-2 variant infection in Santiago, Chile. Front. Public Health 2022, 10, 984433. doi: 10.3389/fpubh.2022.984433.

91. Thornycroft, P.; Brown, W. South African doctor who raised alarm about omicron variant says symptoms are ‘unusual but mild’. Telegraph 27 November 2021. Available at: https://www.telegraph.co.uk/global-health/science-and-disease/south-african-doctor-raised-alarm-omicron-variant-says-symptoms/ xAccessed 30 November 2022.

92. Rashid, N. Sars-CoV-2 B.1.1.529 (Omicron) variant outbreak: case series presentations and response to treatment at the Islamic University in Uganda health facility. ScienceRise 2022, 1, 36–40. https://doi.org/10.21303/2313-8416.2022.002369

93. Chibwana, M.G.; Thole, H.W.; Anscombe, C.; Ashton, P.M.; Green, E.; Barnes, K.G.; Cornick, J.; Turner, A.; Witte, D.; Nthala, S.; et al. Differential symptoms among COVID-19 outpatients before and during periods of SARS-CoV-2 Omicron variant dominance in Blantyre, Malawi: a prospective observational study. [Preprint] medRxiv posted on July 17, 2022. doi: https://doi.org/10.1101/2022.07.15.22277665

94. Mndala, L.; Monk, E.J.M.; Phiri, D.; Riches, J.; Makuluni, R.; Gadama, L.; Kachale, F.; Bilesi, R.; Mbewe, M.; Likaka, A.; et al. Comparison of maternal and neonatal outcomes of COVID-19 before and after SARS-CoV-2 omicron emergence in maternity facilities in Malawi (MATSurvey): data from a national maternal surveillance platform. Lancet Glob. Health 2022, 10, e1623–e1631. doi: 10.1016/S2214-109X(22)00359-X.

95. Moolla, M.S.; Maponga, T.; Moolla, H.; Kollenberg, E.; Anie, S.; Moolla, A.; Moodley, D.; Lalla, U.; Allwood, B.W.; Schrueder, N.; et al. A tale of two waves: Characteristics and outcomes of COVID-19 admissions during the omicron-driven 4th wave in Cape Town, South Africa and implications for the future. IJID Reg. 2022 Nov 24. doi: 10.1016/j.ijregi.2022.11.008. Epub ahead of print.

96. Hajjo, R.; AbuAlSamen, M.M.; Alzoubi, H.M.; Alqutob, R. The Epidemiology of Hundreds of Individuals Infected with Omicron BA.1 in Middle Eastern Jordan. medRxiv [Preprint] Jan 25, 2022. doi: 10.1101/2022.01.23.22269442.

97. Akavian, I.; Nitzan, I.; Talmy, T.; Nitecki, M.; Gendler, S.; Besor, O. SARS-CoV-2 Omicron Variant: Clinical Presentation and Occupational Implications in Young and Healthy IDF Soldiers. Mil. Med. 2022 Sep 3:usac263. doi: 10.1093/milmed/usac263.

98. Kirca, F.; Aydoğan, S.; Gözalan, A.; Kayipmaz, A.E.; Özdemir, F.A.E.; Tekçe, Y.T.; Beşer, İ.O.; Gün, P.; Ökten, R.S.; Dinç, B. Comparison of clinical characteristics of wild-type SARS-CoV-2 and Omicron. Rev. Assoc. Med. Bras. (1992). 2022, 68, 1476–1480. doi: 10.1590/1806-9282.20220880.

99. von Bartheld, C.S.; Hagen, M.M.; Butowt, R. Prevalence of Chemosensory Dysfunction in COVID-19 Patients: A Systematic Review and Meta-analysis Reveals Significant Ethnic Differences. ACS Chem. Neurosci. 2020, 11, 2944–2961. doi: 10.1021/acschemneuro.0c00460.

100. Mutiawati, E.; Fahriani, M.; Mamada, S.S.; Fajar, J.K.; Frediansyah, A.; Maliga, H.A.; Ilmawan, M.; Emran, T.B.; Ophinni, Y.; Ichsan, I.; et al. Anosmia and dysgeusia in SARS-CoV-2 infection: incidence and effects on COVID-19 severity and mortality, and the possible pathobiology mechanisms - a systematic review and meta-analysis. F1000Res. 2021, 10, 40. doi: 10.12688/f1000research.28393.1.

101. Kumar, S.; Thambiraja, T.S.; Karuppanan, K.; Subramaniam, G. Omicron and Delta variant of SARS-CoV-2: A comparative computational study of spike protein. J. Med. Virol. 2022, 94, 1641–1649. doi: 10.1002/jmv.27526.

102. Butowt, R.; Bilinska, K.; von Bartheld, C.S. Olfactory dysfunction in COVID-19: new insights into the underlying mechanisms. Trends Neurosci. 2023, 46, 75–90. doi: 10.1016/j.tins.2022.11.003.

103. Jackson, C.B.; Farzan, M.; Chen, B.; Choe, H. Mechanisms of SARS-CoV-2 entry into cells. Nat. Rev. Mol. Cell. Biol. 2022, 23, 3–20. doi: 10.1038/s41580-021-00418-x.

104. Bilinska, K.; Jakubowska, P.; von Bartheld, C.S.; Butowt, R. Expression of the SARS-CoV-2 Entry Proteins, ACE2 and TMPRSS2, in Cells of the Olfactory Epithelium: Identification of Cell Types and Trends with Age. ACS Chem. Neurosci. 2020, 11, 1555–1562. doi: 10.1021/acschemneuro.0c00210.

105. Brann, D.H.; Tsukahara, T.; Weinreb, C.; Lipovsek, M.; Van den Berge, K.; Gong, B.; Chance, R.; Macaulay, I.C.; Chou, H.J.; Fletcher, R.B.; et al. Non-neuronal expression of SARS-CoV-2 entry genes in the olfactory system suggests mechanisms underlying COVID-19-associated anosmia. Sci. Adv. 2020, 6, eabc5801. doi: 10.1126/sciadv.abc5801.

106. Karimian, A.; Behjati, M.; Karimian, M. Molecular mechanisms involved in anosmia induced by SARS-CoV-2, with a focus on the transmembrane serine protease TMPRSS2. Arch. Virol. 2022, 167, 1931–1946. doi: 10.1007/s00705-022-05545-0.

107. Majdoul, S.; Compton, A.A. Lessons in self-defence: inhibition of virus entry by intrinsic immunity. Nat. Rev. Immunol. 2022, 22, 339–352. doi: 10.1038/s41577-021-00626-8.

108. Zazhytska, M.; Kodra, A.; Hoagland, D.A.; Frere, J.; Fullard, J.F.; Shayya, H.; McArthur, N.G.; Moeller, R.; Uhl, S.; Omer, A.D.; et al. Non-cell-autonomous disruption of nuclear architecture as a potential cause of COVID-19-induced anosmia. Cell 2022, 185, 1052-1064.e12. doi: 10.1016/j.cell.2022.01.024.

109. Armando, F.; Beythien, G.; Kaiser, F.K.; Allnoch, L.; Heydemann, L.; Rosiak, M.; Becker, S.; Gonzalez-Hernandez, M.; Lamers, M.M.; Haagmans, B.L.; et al. SARS-CoV-2 Omicron variant causes mild pathology in the upper and lower respiratory tract of hamsters. Nat. Commun. 2022, 13, 3519. doi: 10.1038/s41467-022-31200-y.

110. Leclerc, S.; Heydel, J.M.; Amossé, V.; Gradinaru, D.; Cattarelli, M.; Artur, Y.; Goudonnet, H.; Magdalou, J.; Netter, P.; Pelczar, H.; et al. Glucuronidation of odorant molecules in the rat olfactory system: activity, expression and age-linked modifications of UDP-glucuronosyltransferase isoforms, UGT1A6 and UGT2A1, and relation to mitral cell activity. Brain Res. Mol. Brain Res. 2002, 107, 201–213. doi: 10.1016/s0169-328x(02)00455-2.

111. Buckley, D.B.; Klaassen, C.D. Tissue- and gender-specific mRNA expression of UDP-glucuronosyltransferases (UGTs) in mice. Drug Metab. Dispos. 2007, 35, 121–127. doi:10.1124/dmd.106.012070

112. Gözen, E.D.; Aliyeva, C.; Tevetoğlu, F.; Karaali, R.; Balkan, İ.İ.; Yener, H.M.; Özdoğan, H.A. Evaluation of Olfactory Function with Objective Tests in COVID-19-Positive Patients: A Cross-Sectional Study. Ear Nose Throat J. 2021, 100, 169S–173S. doi: 10.1177/0145561320975510.

113. Lima, M.A.; Silva, M.T.T.; Oliveira, R.V.; Soares, C.N.; Takano, C.L.; Azevedo, A.E.; Moraes, R.L.; Rezende, R.B.; Chagas, I.T.; Espíndola, O.; et al. Smell dysfunction in COVID-19 patients: More than a yes-no question. J. Neurol. Sci. 2020, 418, 117107. doi: 10.1016/j.jns.2020.117107.

114. Jensen, M.M.; Larsen, K.D.; Homøe, A.S.; Simonsen, A.L.; Arndal, E.; Koch, A.; Samuelsen, G.B.; Nielsen, X.C.; Todsen, T.; Homøe, P. Subjective and psychophysical olfactory and gustatory dysfunction among COVID-19 outpatients; short- and long-term results. PLoS One 2022, 17, e0275518. doi: 10.1371/journal.pone.0275518.

115. Kaya, A.; Altıparmak, S.; Yaşar, M.; Özcan, İ.; Çelik, İ. Objective Evaluation of Smell and Taste Senses in COVID-19 Patients. Turk. Arch. Otorhinolaryngol. 2022, 60, 128–133. doi: 10.4274/tao.2022.2022-6-1.

116. Lechien, J.R.; Cabaraux, P.; Chiesa-Estomba, C.M.; Khalife, M.; Hans, S.; Calvo-Henriquez, C.; Martiny, D.; Journe, F.; Sowerby, L.; Saussez, S. Objective olfactory evaluation of self-reported loss of smell in a case series of 86 COVID-19 patients. Head Neck 2020, 42, 1583–1590. doi: 10.1002/hed.26279.

117. Lechien, J.R.; Chiesa-Estomba, C.M.; Hans, S.; Barillari, M.R.; Jouffe, L.; Saussez, S. Loss of Smell and Taste in 2013 European Patients with Mild to Moderate COVID-19. Ann. Intern. Med. 2020, 173, 672–675. doi: 10.7326/M20-2428.

118. Romero-Gameros, C.A.; Waizel-Haiat, S.; Mendoza-Zubieta, V.; Anaya-Dyck, A.; López-Moreno, M.A.; Colin-Martinez, T.; Martínez-Ordaz, J.L.; Ferat-Osorio, E.; Vivar-Acevedo, E.; Vargas-Ortega, G.; et al. Evaluation of predictive value of olfactory dysfunction, as a screening tool for COVID-19. Laryngoscope Investig. Otolaryngol. 2020, 5, 983–991. doi: 10.1002/lio2.482.

119. Hintschich, C.A.; Wenzel, J.J.; Hummel, T.; Hankir, M.K.; Kühnel, T.; Vielsmeier, V.; Bohr, C. Psychophysical tests reveal impaired olfaction but preserved gustation in COVID-19 patients. Int. Forum Allergy Rhinol. 2020, 10, 1105–1107. doi: 10.1002/alr.22655.

120. Le Bon, S.D.; Pisarski, N.; Verbeke, J.; Prunier, L.; Cavelier, G.; Thill, M.P.; Rodriguez, A.; Dequanter, D.; Lechien, J.R.; Le Bon, O.; et al. Psychophysical evaluation of chemosensory functions 5 weeks after olfactory loss due to COVID-19: a prospective cohort study on 72 patients. Eur. Arch Otorhinolaryngol. 2021, 278, 101–108. doi: 10.1007/s00405-020-06267-2.

121. Hummel, T.; Whitcroft, K.L.; Andrews, P.; Altundag, A.; Cinghi, C.; Costanzo, R.M.; Damm, M.; Frasnelli, J.; Gudziol, H.; Gupta, N.; et al. Position paper on olfactory dysfunction. Rhinol. Suppl. 2017, 54, 1–30. doi: 10.4193/Rhino16.248.

122. Fahmy, M.; Whitcroft, K. Psychophysical Testing in Chemosensory Disorders. Curr Otorhinolaryngol. Rep. 2022, 10, 393–404. doi: 10.1007/s40136-022-00429-y.

123. Killingley, B.; Mann, A.J.; Kalinova, M.; Boyers, A.; Goonawardane, N.; Zhou, J.; Lindsell, K.; Hare, S.S.; Brown, J.; Frise, R.; et al. Safety, tolerability and viral kinetics during SARS-CoV-2 human challenge in young adults. Nat. Med. 2022, 28, 1031–1041. doi: 10.1038/s41591-022-01780-9.

124. Mariño-Sánchez, F.; Santamaría-Gadea, A.; de Los Santos, G.; Alobid, I.; Mullol, J. Psychophysical olfactory testing in COVID-19: is smell function really impaired in nearly all patients? Int. Forum Allergy Rhinol. 2020, 10, 951–952. doi: 10.1002/alr.22639.

125. Desiato, V.M.; Levy, D.A.; Byun, Y.J.; Nguyen, S.A.; Soler, Z.M.; Schlosser, R.J. The Prevalence of Olfactory Dysfunction in the General Population: A Systematic Review and Meta-analysis. Am. J. Rhinol. Allergy. 2021, 35, 195–205. doi: 10.1177/1945892420946254.

126. Agyeman, A.A.; Chin, K.L.; Landersdorfer, C.B.; Liew, D.; Ofori-Asenso, R. Smell and Taste Dysfunction in Patients With COVID-19: A Systematic Review and Meta-analysis. Mayo Clin. Proc. 2020, 95, 1621–1631. doi: 10.1016/j.mayocp.2020.05.030.

127. Wang, X.; Chang, H.; Tian, H.; Zhu, Y.; Li, J.; Wei, Z.; Wang, Y.; Xia, A.; Ge, Y.; Liu, G.; et al. Epidemiological and clinical features of SARS-CoV-2 infection in children during the outbreak of Omicron variant in Shanghai, March 7-31, 2022. Influenza Other Respir. Viruses 2022, 16, 1059–1065. doi: 10.1111/irv.13044.

128. Lee, B.R.; Harrison, C.J.; Myers, A.L.; Jackson, M.A.; Selvarangan, R. Differences in pediatric SARS-CoV-2 symptomology and Co-infection rates among COVID-19 Pandemic waves. J. Clin. Virol. 2022, 154, 105220. doi: 10.1016/j.jcv.2022.105220.

129. Shoji, K.; Akiyama, T.; Tsuzuki, S.; Matsunaga, N.; Asai, Y.; Suzuki, S.; Iwamoto, N.; Funaki, T.; Ohmagari, N. Clinical characteristics of COVID-19 in hospitalized children during the Omicron variant predominant period. J. Infect. Chemother. 2022, 28, 1531–1535. doi: 10.1016/j.jiac.2022.08.004.

130. Stopyra, L.; Kowalik, A.; Stala, J.; Majchrzak, I.; Szebla, J.; Jakosz, M.; Grzywaczewska, K.; Kwinta, P. Characteristics of Hospitalized Pediatric Patients in the First Five Waves of the COVID-19 Pandemic in a Single Center in Poland-1407 Cases. J. Clin. Med. 2022, 11, 6806. doi: 10.3390/jcm11226806.

131. Meyer, M.; Ruebsteck, E.; Dewald, F.; Klein, F.; Lehmann, C.; Huenseler, C.; Weber, L.T. Clinical Aspects of the Subsequent SARS-CoV-2 Waves in Children from 2020 to 2022-Data from a Local Cohort in Cologne, Germany (n = 21,635). Viruses 2022, 14, 1607. doi: 10.3390/v14081607.

132. Smith, H.; Mahon, A.; Moss, A.; Rao, S. SARS-CoV-2 infection in children evaluated in an ambulatory setting during Delta and Omicron time periods. J. Med. Virol. 2023, 95, e28318. doi: 10.1002/jmv.28318.

133. Pieniak, M.; Oleszkiewicz, A.; Avaro, V.; Calegari, F.; Hummel, T. Olfactory training - Thirteen years of research reviewed. Neurosci. Biobehav. Rev. 2022, 141, 104853. doi: 10.1016/j.neubiorev.2022.104853.

134. Lechien, J.R.; Chiesa-Estomba, C.M.; Radulesco, T.; Michel, J.; Vaira, L.A.; Le Bon, S.D.; Horoi, M.; Falanga, C.; Barillari, M.R.; Hans, S.; et al. Clinical features of patients who had two COVID-19 episodes: a European multicentre case series. J. Intern. Med. 2021, 290, 421–429. doi: 10.1111/joim.13259.

135. Lechien, J.R.; Chiesa-Estomba, C.M.; Vaira, L.A.; Saussez, S.; Hans, S. COVID-19 Reinfection and Second Episodes of Olfactory and Gustatory Dysfunctions: Report of First Cases. Ear Nose Throat J. 2022, 101, 499–500. doi: 10.1177/0145561320970105.

136. Boulware, D.R.; Murray, T.A.; Proper, J.L.; Tignanelli, C.J.; Buse, J.B.; Liebovitz, D.M.; Nicklas, J.M.; Cohen, K.; Puskarich, M.A.; Belani, H.K.; et al. Impact of SARS-CoV-2 vaccination and booster on COVID-19 symptom severity over time in the COVID-OUT trial. Clin. Infect. Dis. 2022 Sep 17:ciac772. Epub ahead of print. doi: 10.1093/cid/ciac772.

